# COVID-19 relevant genetic variants confirmed in an admixed population

**DOI:** 10.1101/2022.04.15.22273925

**Authors:** Tomas Texis, José Luis Cruz-Jaramilllo, Willebaldo García-Muñoz, Lourdes Anzures-Cortés, Lorenza Haddad-Talancón, Sergio Sánchez-García, María del Carmen Jiménez Martínez, Edgar Pérez Barragán, Alejandro Nieto-Patlán, José D. Martínez-Ezquerro, Kenneth Rubio-Carrasco, Mauricio Rodríguez-Dorantes, Sergio Cortés-Ramírez, Gabriela Mellado-Sánchez, Sonia Mayra Pérez-Tapia, Vanessa Gonzalez-Covarrubias

## Abstract

The dissection of factors that contribute to COVID-19 infection and severity has overwhelmed the scientific community for almost 2 years. Current reports highlight the role of in disease incidence, progression, and severity. Here, we aimed to confirm the presence of previously reported genetic variants in an admixed population. Allele frequencies were assessed and compared between the general population (N=3079) for which at least 30% have not been infected with SARS-CoV2 as per July 2021 versus COVID-19 patients (N=106).

Genotyping data from the Illumina GSA array was used to impute genetic variation for 14 COVID-relevant genes, using the 1000G phase 3 as reference based on the human genome assembly hg19, following current standard protocols and recommendations for genetic imputation. Bioinformatic and statistical analyses were performed using *MACH v1.0*, R, and PLINK.

A total of 7953 variants were imputed on, *ABO, CCR2, CCR9, CXCR6, DPP9, FYCO1, IL10RB/IFNAR2, LZTFL1, OAS1, OAS2, OAS3, SLC6A20, TYK2*, and *XCR1*. Statistically significant allele differences were reported for 10 and 7 previously identified and confirmed variants, *ABO* rs657152, *DPP9* rs2109069, *LZTFL1* rs11385942, *OAS1* rs10774671, *OAS1* rs2660, *OAS2* rs1293767, and *OAS3* rs1859330 p<0.03. In addition, we identified 842 variants in these COVID-related genes with significant allele frequency differences between COVID patients and the general population (p-value <E-2 – E-179).

Our observations confirm the presence of genetic differences in COVID-19 patients in an admixed population and prompts for the investigation of the statistical relevance of additional variants on these and other genes that could identify local and geographical patterns of COVID-19.

## Introduction

Most COVID cases are mild, but between 10 – 15% of patients may develop a severe disease with or without metabolic comorbidities (Havervall et al., 2022). For almost two years the scientific community has been trying to depict all risk factors that convey to COVID-19 infection, susceptibility, respiratory failure, and death (Asgari & Pousaz, 2021; Butler-Laporte et al., 2021; Freuer, Linseisen, & Meisinger, 2021; Leong et al., 2021; Wendt et al., 2021). Initial reports of the COVID-19 pandemic identified male and older age as risk determinants for increased severity, respiratory stress, and death (Santesmasses et al., 2020; Zhu et al., 2020). The aging of the adaptive and innate immune system can explain, in part the cytokine storm and the inability of the immune system to efficiently fight against SARS-CoV-2 (Mueller, Mcnamara, & Sinclair, 2020). The investigation of human genetic susceptibility quickly prompted the creation of collaborative research groups such as The Sex, Gender and COVID-19 Project, that attempted to explain why 20% of males were more likely to be hospitalized and then die compared to women. T. Takahashi et al. reported that females show a more robust T cell activation while immune cytokines, associated with a worse progression, are higher in males (Takahashi et al., 2020). Other hypotheses revolve around variation on the X chromosome such as a “XXX” hypercoagulation karyotype and genes, *TLR7* and *ACE2* the latter coding for the SARS-CoV-2 receptor (Li, Jerkic, Slutsky, & Zhang, 2020; Tsiambas et al., 2020; Tümer, Kiliçaslan, & Akinci, 2021). Endeavors of the COVID-19 Host Genetics Initiative (www.covid19hg.org), lead several GWAs investigations and the identification of over a dozen gene variants associated to COVID susceptibility and severity, some of which were identified only in individuals with East Asian ancestry (Asgari & Pousaz, 2021). In September 2020 the UK based Genetics of Mortality in Critical Care (GenOMICC) group identified and replicated at least 9 variants associated with critical COVID-19 in 2244 unrelated patients on genes, *IFNAR2, HLA-G, CCR2, OAS1-3, DPP9, TYK2*, highlighting the host antiviral and inflammation mechanisms in the development of critical illness (Pairo-Castineira et al., 2020a). Soon after, in October 2020 the Severe COVID-19 GWAS Group studied 1980 COVID patients with respiratory failure identifying two main gene clusters on chromosomes 3 and 9, covering several loci and at least 10 genes including, *SLC6A20, LZTFL1, CCR9, FYCO1, CXCR6, ABO*, and *XCR1* (K Niemi et al., 2021). Almost a year later The COVID-19 Host Genetics Initiative performed genetic meta-analyses to pinpoint variation that may lead to biological insights on disease onset, progression, severity, and to identify therapeutic mechanistic targets (K Niemi et al., 2021). In their report the authors confirmed previous research and prioritized gene variants according to their plausible role as causation/severity of COVID-19. These included six loci with protein alterations and in LD with lead variants, *TYK2* rs74956615 previously reported to protect against autoimmune diseases, *PPP1R15A* rs11541192 strongly associated with SARS-CoV-2 infection, and *IFNAR2/IL10RB, OAS1-3, FOXP4*, and *ABO* variants that may potentially alter lung gene expression. This report also highlighted the fact that most studies have included patients with European ancestry (K Niemi et al., 2021). Thereafter, results from GWAS motivated the development of models to predict the risk of severe COVID-19 and in March 2021, G. Dite et. al. published a validated model to assess the risk of COVID-19 for the UK Biobank participants. The model included genetic variants from previous research (Dite, Murphy, & Allman, 2021a), new variation from the COVID-19 Host Genetics Initiative, and variants from P. Castineira et al. (Pairo-Castineira et al., 2020b). The final prediction model included clinical variants (age, sex, BMI, Caucasian/other, cerebrovascular disease, diabetes, hypertension, hematological cancer, cancer, kidney, and respiratory disease) and seven SNVs of the 116 initially tested (rs112641600, rs10755709, 118072448, rs7027911, rs71481792, rs112317747, rs2034831) all on genes related to infection and immunity (Dite, Murphy, & Allman, 2021b), but the utility and replication of this model in other populations remains to be investigated. A recent review on the genetic susceptibility and response to COVID-19 summarizes genetic associations reported up to date for 27 studies listing 81 variants, most of them identified in a few ethnic groups. The World Health Organization reported in December 2021, 290 million COVID-19 cases and 5.45 million related deaths world-wide. For the former Mexico ranked 15^th^ and for the latter 4^th^, which highlights the relevance of comorbidities, and local and geographical determinants impacting a mortality rate which does not parallel in proportion the infection rate. It is also possible that infection statistics not fully depict a country’s reality since COVID-19 testing has not been equally deployed in all countries. According to the Mexican government and graphic-reuters.com, by the end of 2021, Mexico has administered 148,938,454 doses of COVID vaccines, enough to have vaccinated about 58% of the country’s population. Moreover, COVID severity is directly related to cardiovascular comorbidities, obesity, and diabetes all showing a high incidence and prevalence among Mexicans. Metabolic and genetic variation research will likely provide with the tools to assess COVID-19 risk considering geographical differences and highly prevalent comorbidities as the model by G. Dite (Dite et al., 2021a, 2021b).

Here, we sought to investigate allele and genotype frequencies of COVID-19 relevant variants in Mexican Mestizos according to published GWAs studies that were available at the onset of this investigation (Pairo-Castineira et al., 2020b; The Severe Covid-19 GWAS Group, 2020). We also compared these allele frequencies (AF) between the general population and COVID-19 patients, and although the sample size of COVID-19 patients was small we were able to confirm previous observations for variants on *ABO* and *OAS1*-*3* and to depict additional allele frequencies differences between groups in COVID related genes. Our study may aid to pinpoint genetic variation that would potentially assess a genetic risk for COVID-19 in an admixed population.

## Methods

### Study participants

One hundred and six COVID-19 patients were recruited by three different institutions, Hospital General de Zona no. 48, Instituto Politecnico Nacional Departamento de Inmunologia, Escuela Nacional de Ciencias Biologicas, Instituto Politecnico Nacional, ENCB-IPN, and Código 46 during 2020. All 3185 participants signed an informed consent, followed the principles of the declaration of Helsinki. The protocol was approved by the Committees of Research, Ethics and Biosafety at Instituto Mexicano del Seguro Social (IMSS) R2018-785-004 and INMEGEN CEI2017/04 & 23/2016/I.

COVID-19, diagnosis in 106 patients was determined by a nasal PCR test for SARS-CoV-2. For the rest of the 3079 samples, non-COVID infection was confirmed in at least in 880 patients by PCR or an antigen test or with a phone call as a follow up appointment from IMSS Centro Medico Nacional Siglo XXI between February and June 2021 confirming negativity for any SARS-CoV-2 test.

For all other participants (N=2199) data were collected from prior and current research protocols and from different collaborating laboratories in Mexico City, for these, 30% of samples were confirmed as non-COVID-19, the remaining cannot be ruled out as COVID or non-COVID but served as a genetic pool to assess allele frequencies of relevant COVID-19 variants in the Mexican population. Therefore, in this study we performed allele frequency comparisons between confirmed COVID-19 patients (N=106) and the general population (N=3079) which included non-COVID confirmed by an antigen or PCR SARS-CoV-2 test (N=880) and individuals unknown diagnosis (N=2199). Comparisons were also performed for COVID-19 (N=106) vs. confirmed non-COVID (N=880) with similar results, but the sample size was substantially reduced and did not benefit the reporting of population allele frequencies.

### Genotyping, imputation, and statistical analyses

For COVID-19 positive patients, DNA was extracted using a commercial kit followed by genotyping using the GSA microarray v.1.0 (Illumina) followed by bioinformatic and genotyping controls. For the rest of the population, genetic data was retrieved from previously genotyping using the same array. The genes selected for consideration for these analyses came from two high impact sources, The Severe COVID-19 GWAS Group (*SLC6A20, LZTFL1, CCR9, FYCO1, CXCR6, XCR1, ABO*) (The Severe Covid-19 GWAS Group, 2020), and the report by Pairo-Castineira et al. (*OAS1, OAS2, OAS3, TYK2, DPP9, IL10RB/IFNAR2, CCR2*) (K Niemi et al., 2021). The GSA Illumina genotyping array can detect 670K common single nucleotide variants (SNVs) but most of the COVID-19 reported loci are not included in this platform and needed imputation. We imputed all variants on these loci/genes according to standard protocols and recommendations for genetic imputation (Marino et al., 2021; Uffelmann et al., 2021) using public data from the 1000G phase 3 as reference based on the human genome assembly hg19 and utilizing current imputation practices with the software *MACH v.1.0* (Auton et al., 2015; Lam, Awasthi, Watson, …, & 2020, n.d.; Liu, Li, Wang, & Li, 2013). Variants that passed bioinformatic quality controls were considered for bioinformatic, allele frequency (AF) comparisons, and statistical analyses (Purcell et al., 2007). We were able to impute and detect in this study group most of the variants previously reported associated to COVID-19. Group comparisons were performed for all variants with allele and genotyping frequencies detected, AF>0.0001 including the assessment of differences in AF between COVID-19 patients (N=106) and the general population (N=3019) i.e. as described before, we did not include a control group, instead we defined a general population group for which about a third were confirmed as never being infected with SARS-CoV-2 as per September 2021. AF statistical differences were assessed using a Chi-square test considering a threshold of P<0.05, calculations were computed using the Software R v4.0.5. Alternatively, comparisons were also performed exclusively for COVID vs. non-COVID with similar results, but the sample size was significantly reduced and did not benefit the reporting of population allele frequencies. Supplemental Table 1 lists all variants identified on these genes with an allele frequency ≥0.01%. Raw imputation data are available from the corresponding author upon reasonable request.

## Results

We report allele and genotype frequencies of all variants detected in genomic regions previously associated with COVID-19 (total SNVs=7953. Supplemental Table 1 (Pairo-Castineira et al., 2020b; The Severe Covid-19 GWAS Group, 2020). Next, we compared allele and genotype frequencies between COVID-19 patients and the general population for all 7953 detected variants with a particular focus on those already consistently reported for COVID-19 patients.

The group here defined as the general population were on average 58 years of age, showed a BMI of 33 kg/m^2^, 68% were diabetic and 48% hypertensive. The COVID-19 group were on average age 42 years old, 50% were diabetic and 70% hypertensive, 94% of them had a confirmed COVID-19 diagnosis based on a PCR or antigen test. Clinical information for BMI was not available for COVID-19 patients and data for T2D and hypertension has not been completely reported (Table 1). Fourteen COVID-19 patients showed severe COVID and required hospitalization, sixteen showed clinically treated moderate symptoms and ten showed mild symptoms, for the remaining patients the medical record has not been completed yet.

**Table 1.** Population characteristics

|  | COVID | Non-COVID |
| --- | --- | --- |
| N | 56 | 2677 |
| Females | 22 | 1529 |
| Age, y, mean (range) | 42 (17 - 73) | 58 (17 - 97) |
| BMI<sup>1</sup> kg/m<sup>2</sup> | 28 (20 – 31) | 33 (17 - 55) |
| T2D<sup>1</sup> | 80% | 1832 |
| Hypertension<sup>1</sup> | 70% | 1272 |
| Hospitalized | 14 | none |
| PCR COVID test | 24 | unknown |
| Other COVID test | 29 | 600 |
| N, severe COVID | 16 | NA |
| N, average symptoms | 23 | NA |
| N, asymptomatic | 16 | NA |

<sup>1</sup> Information for BMI and T2D was recorded for ~50% of the COVID patients, i.e., about 27 out of 56 patients were clinically known to have diabetes and reported a BMI value, for the other half of the COVID patients, data were not available.

We imputed 7953 SNVs on 14 genes/loci on chromosomes 3, 9, 12, 19, and 21 with variation that has been consistently associated to COVID-19 in the recent literature (Aydillo, Esther Babady, & Kamboj, 2020a; Initiative, 2020; Kaser, 2020). These loci include COVID-19 related genes, *ABO, CCR2, CCR9, CXCR6, DPP9, FYCO1, IL10RB/IFNAR2, LZTFL1, OAS1, OAS2, OAS3, SLC6A20, TYK2*, and *XCR1*. Of the 7953 SNVs imputed, 3696 were detected in Mexican Mestizos i.e., variants with at least one alternative allele. Common variation i.e., an allele frequency > 0.01 was observed for 2645 variants in both study groups and most SNVs showed allele frequency of: mean:0.0663, median 0.002, range 0.00016-0.500, (Supplemental Table 1).

When comparing AF between COVID-19 samples versus the general population (N=106 vs. 3019), we confirmed the statistical significance of 7 out of 10 variants that have been previously associated to COVID-19 infection and severe disease, rs657152, rs1859330, rs1293767, rs10774671, rs2660, rs2109069, rs11385942. Variants, rs8176747, rs2285932, and rs2236757, did not show relevant allele frequency differences in our group comparisons despite being reported as COVID-19 related, i.e., seven of ten variants previously associated to COVID-19 were confirmed in this study showing significant AF differences in COVID-19 patients versus the general population (Table 2). Additional to the comparisons on previously reported variants, we identified 13 SNVs on these regions with apparent and significant AF differences between COVID-19 patients and the general population. The most relevant differences were found on *CXCR6, FYCO1, NRBF2P2, XCR1, ABO, OAS3*, and *DPP9* for the latter we observed linkage disequilibrium on rs72977997 and rs118149076, (Table 2) all variants and AF can be found in Supplemental Table 1.

**Table 2.**
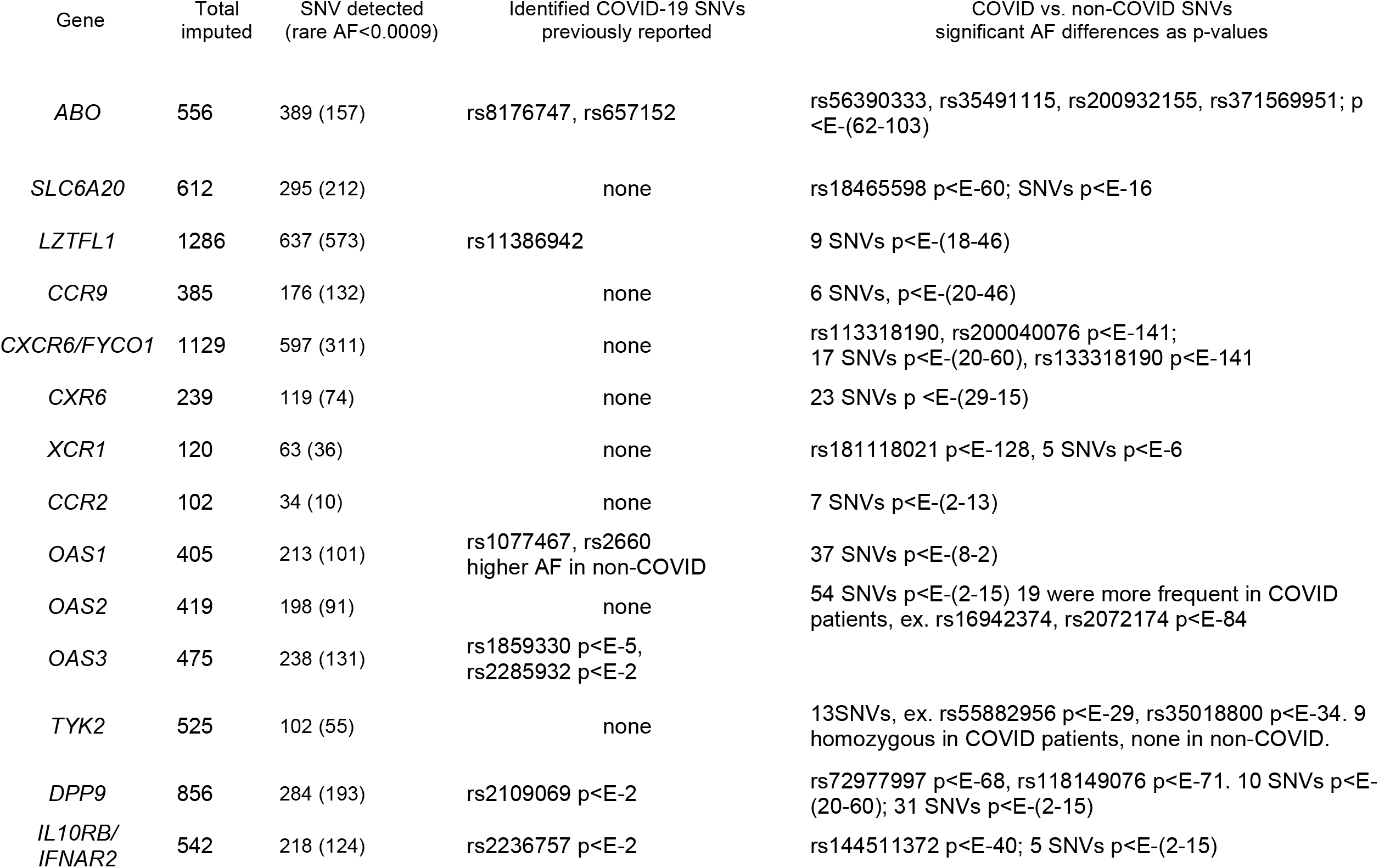
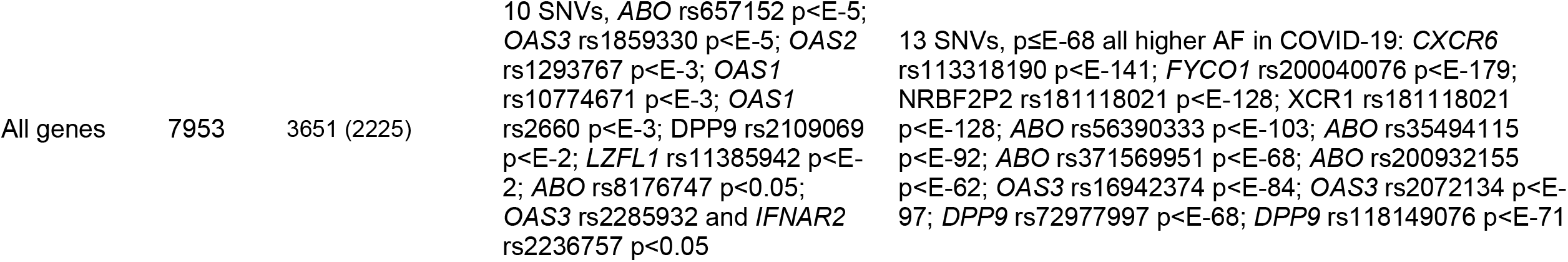
Summary of genetic variants (SNV) in COVID-19 associated genes

### Genetic variation exclusive of COVID-19 samples

Interestingly 43 variants showed an AF of zero in the general population (N=3079) but were present in COVID patients i.e., 43 variants showed the reference or “wild type” genotype in the general population but were homozygous or heterozygous in COVID-19 samples. Of these *ABO* rs56390333 (AF=0.076) and *FYCO1* rs200040076 (AF=0.132) showed the most significant differences in AF with 5 and 11 with a homozygous genotype in COVID-19 samples respectively (p-value=E-179, and E-62, Table 2). Other genes that also showed variation exclusively in COVID patients were, *CCR9* (9 SNVs), *OAS2* (3 SNVs), *LZTFL1* (12 SNVs), *SLC6A20* (8 SNVs), *IFNAR2, DPP9*, and *IL10RB*, the latter three with one SNV each, statistical significance ranged from p-value < E-8 to E-46 (Supplemental Table 1).

### Common variation in COVID-19 and the general population

Next, we compared allele and genotype frequencies between COVID-19 patients and the general population for all variants with an AF>0, Table 3 lists variants with the largest statistical differences. Of the several loci clustered on chromosomes, 19, 21, 3, and 9 consistently associated with COVID-19 (Aydillo et al., 2020a; Pairo-Castineira et al., 2020b; Yildirim, Sahin, Yazar, & Bozok Cetintas, 2021) we identified 10 of these loci in Mexican Mestizos, seven of them showing significant AF differences between COVID patients and the general population including, *ABO* rs657152 p-value=7.2E-05, *DPP9* rs2109069 p-value=0.014, *LZTFL1* rs11385942 p-value=0.029, *OAS1* rs10774671 p-value=0.0035, *OAS1* rs2660 p-value=0.007, *OAS2* rs1293767 p-value=0.00501, and *OAS3* rs1859330 p-value=2.9E-05. Except for *ABO* rs657152 and *DPP9* rs2109069 the above SNVs did not comply with Hardy-Weinberg equilibrium p<0.003. Reports by the Severe COVID-19 GWAS and the GenOMICC groups included the above-mentioned variants for which we identified AF differences in COVID-19 samples but for the three remaining of the 10 sought loci here, *ABO* rs8176747, *IFNAR2* rs2236757, *OAS3* rs2285932, we did not observe significant AF differences (Table 3). Supplemental Table 1 includes a full list of genetic variants and AF differences.

**Table 3.**
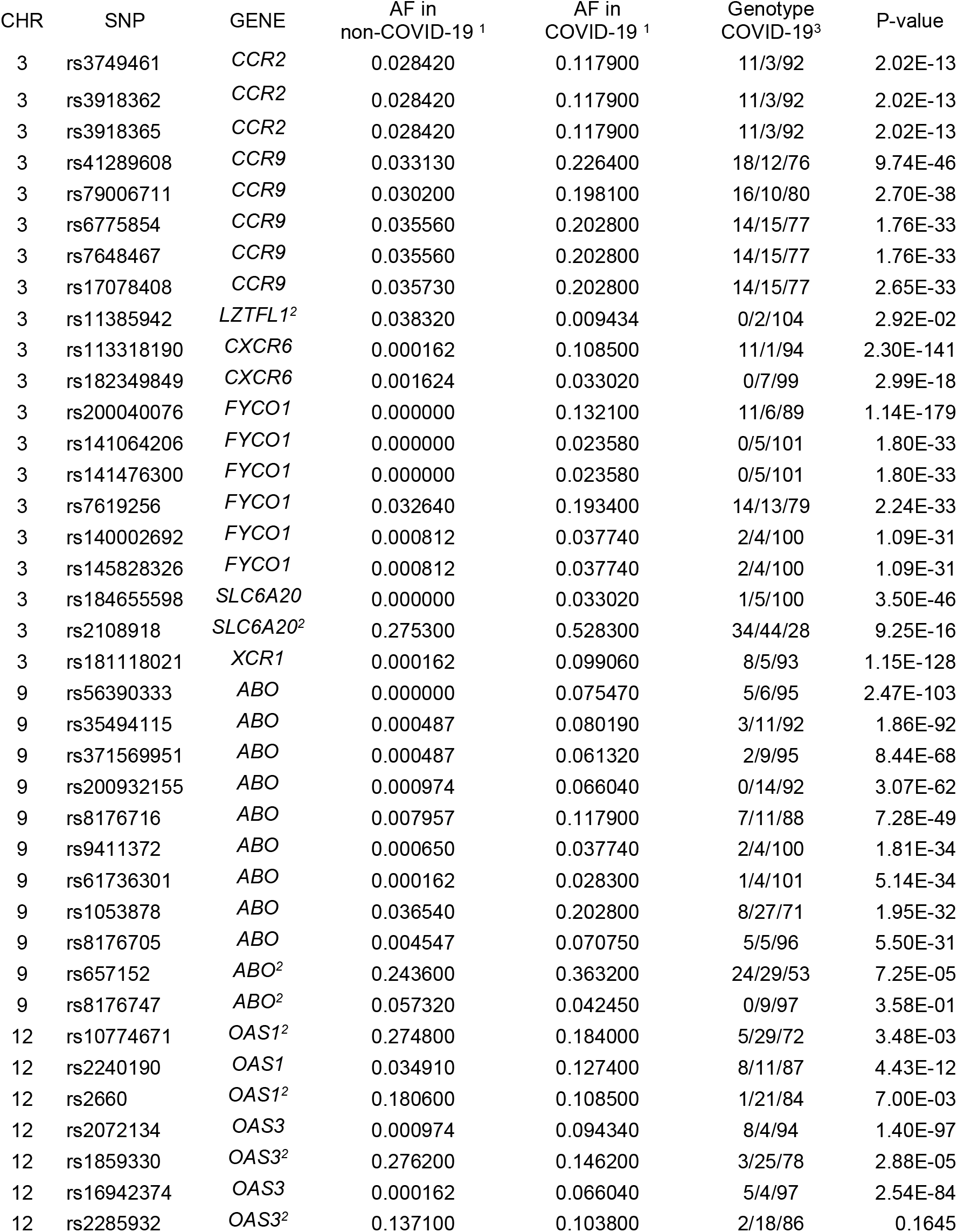
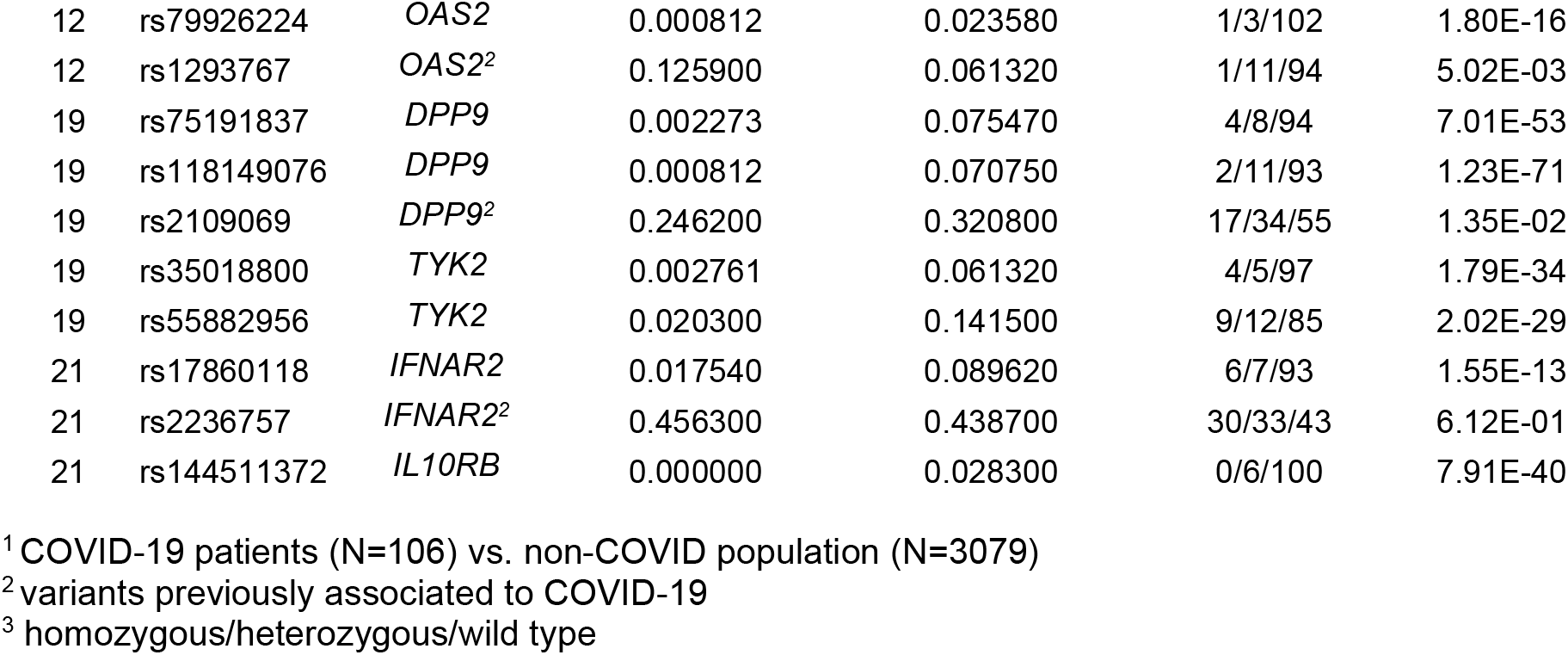
Allele frequency in COVID vs. non-COVID patients in COVID-19 associated variants

| CHR | SNP | GENE | AF in<br>non-COVID-19 <sup>1</sup> | AF in<br>COVID-19 <sup>1</sup> | Genotype<br>COVID-19<sup>3</sup> | P-value |
| --- | --- | --- | --- | --- | --- | --- |
| 3 | rs3749461 | *CCR2* | 0.028420 | 0.117900 | 11/3/92 | 2.02E-13 |
| 3 | rs3918362 | *CCR2* | 0.028420 | 0.117900 | 11/3/92 | 2.02E-13 |
| 3 | rs3918365 | *CCR2* | 0.028420 | 0.117900 | 11/3/92 | 2.02E-13 |
| 3 | rs41289608 | *CCR9* | 0.033130 | 0.226400 | 18/12/76 | 9.74E-46 |
| 3 | rs79006711 | *CCR9* | 0.030200 | 0.198100 | 16/10/80 | 2.70E-38 |
| 3 | rs6775854 | *CCR9* | 0.035560 | 0.202800 | 14/15/77 | 1.76E-33 |
| 3 | rs7648467 | *CCR9* | 0.035560 | 0.202800 | 14/15/77 | 1.76E-33 |
| 3 | rs17078408 | *CCR9* | 0.035730 | 0.202800 | 14/15/77 | 2.65E-33 |
| 3 | rs11385942 | *LZTFL1*<sup>2</sup> | 0.038320 | 0.009434 | 0/2/104 | 2.92E-02 |
| 3 | rs113318190 | *CXCR6* | 0.000162 | 0.108500 | 11/1/94 | 2.30E-141 |
| 3 | rs182349849 | *CXCR6* | 0.001624 | 0.033020 | 0/7/99 | 2.99E-18 |
| 3 | rs200040076 | *FYCO1* | 0.000000 | 0.132100 | 11/6/89 | 1.14E-179 |
| 3 | rs141064206 | *FYCO1* | 0.000000 | 0.023580 | 0/5/101 | 1.80E-33 |
| 3 | rs141476300 | *FYCO1* | 0.000000 | 0.023580 | 0/5/101 | 1.80E-33 |
| 3 | rs7619256 | *FYCO1* | 0.032640 | 0.193400 | 14/13/79 | 2.24E-33 |
| 3 | rs140002692 | *FYCO1* | 0.000812 | 0.037740 | 2/4/100 | 1.09E-31 |
| 3 | rs145828326 | *FYCO1* | 0.000812 | 0.037740 | 2/4/100 | 1.09E-31 |
| 3 | rs184655598 | *SLC6A20* | 0.000000 | 0.033020 | 1/5/100 | 3.50E-46 |
| 3 | rs2108918 | *SLC6A20*<sup>2</sup> | 0.275300 | 0.528300 | 34/44/28 | 9.25E-16 |
| 3 | rs181118021 | *XCR1* | 0.000162 | 0.099060 | 8/5/93 | 1.15E-128 |
| 9 | rs56390333 | *ABO* | 0.000000 | 0.075470 | 5/6/95 | 2.47E-103 |
| 9 | rs35494115 | *ABO* | 0.000487 | 0.080190 | 3/11/92 | 1.86E-92 |
| 9 | rs371569951 | *ABO* | 0.000487 | 0.061320 | 2/9/95 | 8.44E-68 |
| 9 | rs200932155 | *ABO* | 0.000974 | 0.066040 | 0/14/92 | 3.07E-62 |
| 9 | rs8176716 | *ABO* | 0.007957 | 0.117900 | 7/11/88 | 7.28E-49 |
| 9 | rs9411372 | *ABO* | 0.000650 | 0.037740 | 2/4/100 | 1.81E-34 |
| 9 | rs61736301 | *ABO* | 0.000162 | 0.028300 | 1/4/101 | 5.14E-34 |
| 9 | rs1053878 | *ABO* | 0.036540 | 0.202800 | 8/27/71 | 1.95E-32 |
| 9 | rs8176705 | *ABO* | 0.004547 | 0.070750 | 5/5/96 | 5.50E-31 |
| 9 | rs657152 | *ABO*<sup>2</sup> | 0.243600 | 0.363200 | 24/29/53 | 7.25E-05 |
| 9 | rs8176747 | *ABO*<sup>2</sup> | 0.057320 | 0.042450 | 0/9/97 | 3.58E-01 |
| 12 | rs10774671 | *OAS1*<sup>2</sup> | 0.274800 | 0.184000 | 5/29/72 | 3.48E-03 |
| 12 | rs2240190 | *OAS1* | 0.034910 | 0.127400 | 8/11/87 | 4.43E-12 |
| 12 | rs2660 | *OAS1*<sup>2</sup> | 0.180600 | 0.108500 | 1/21/84 | 7.00E-03 |
| 12 | rs2072134 | *OAS3* | 0.000974 | 0.094340 | 8/4/94 | 1.40E-97 |
| 12 | rs1859330 | *OAS3*<sup>2</sup> | 0.276200 | 0.146200 | 3/25/78 | 2.88E-05 |
| 12 | rs16942374 | *OAS3* | 0.000162 | 0.066040 | 5/4/97 | 2.54E-84 |
| 12 | rs2285932 | *OAS3*<sup>2</sup> | 0.137100 | 0.103800 | 2/18/86 | 0.1645 |
| 12 | rs79926224 | *OAS2* | 0.000812 | 0.023580 | 1/3/102 | 1.80E-16 |
| 12 | rs1293767 | *OAS2*<sup>2</sup> | 0.125900 | 0.061320 | 1/11/94 | 5.02E-03 |
| 19 | rs75191837 | *DPP9* | 0.002273 | 0.075470 | 4/8/94 | 7.01E-53 |
| 19 | rs118149076 | *DPP9* | 0.000812 | 0.070750 | 2/11/93 | 1.23E-71 |
| 19 | rs2109069 | *DPP9*<sup>2</sup> | 0.246200 | 0.320800 | 17/34/55 | 1.35E-02 |
| 19 | rs35018800 | *TYK2* | 0.002761 | 0.061320 | 4/5/97 | 1.79E-34 |
| 19 | rs55882956 | *TYK2* | 0.020300 | 0.141500 | 9/12/85 | 2.02E-29 |
| 21 | rs17860118 | *IFNAR2* | 0.017540 | 0.089620 | 6/7/93 | 1.55E-13 |
| 21 | rs2236757 | *IFNAR2*<sup>2</sup> | 0.456300 | 0.438700 | 30/33/43 | 6.12E-01 |
| 21 | rs144511372 | *IL10RB* | 0.000000 | 0.028300 | 0/6/100 | 7.91E-40 |

<sup>1</sup> COVID-19 patients (N=106) vs. non-COVID population (N=3079)
<sup>2</sup> variants previously associated to COVID-19
<sup>3</sup> homozygous/heterozygous/wild type

### AF differences not previously reported

In addition to confirming previous results, we identified over 100 variants with significant AF differences between the general population and COVID-19 patients (Table 3). Our results add to the collection of variants on genes already related to COVID-19. The most relevant were 13 variants, 4 on *ABO*, 3 on *DPP9*, 2 on *FYCO1, OAS3*, and one on *CXCR6* and *XCR1*, all showing the alternative allele >10-fold more frequently in COVID-19 patients compared to the average population (p-value<E-53 – E-179).

### Protective or susceptibility variation

Human genetic loci have been defined as protective or susceptibility variants depending on their allele frequency (higher vs lower) effect size, or odds ratio values on association models towards COVID-19 (Huffman et al., 2022; Velavan et al., 2021; Zeberg & Pääbo, 2021a). Here, we had AF information and could classify variants as higher in frequency of the alternative allele in COVID-19 patients versus the general population. We performed this analysis and observed 287 variants with a statistically significant higher AF in the general population vs. COVID-19 patients, the most significant were *SLC6A20* rs2742396 (AF: 0.445 vs. 0.212, p-value 2E-11), *OAS2* rs1293758 (AF: 0.295 vs. 0.104) and *OAS2* rs1298961 (AF:0.330 vs. 0.0044; p-value 8E-11, in almost complete LD: D=1.0, R=0.962). *OAS3* rs107746 and *OAS1* rs2660 p<E-3, previously reported as protective variants against COVID-19 infection, were also identified here but with a lower statistical significance. All other statistically relevant variants can be found in Supplemental Table 1. In contrast, variants whose AF were higher in COVID-19 patients compared to the general population could be considered as deleterious. In this regard, we observed 734 variants with statistically significant higher allele frequencies in COVID-19 samples (p-value<0.05). The most statistically different were, *CXCR6* rs113318190 (AF: 0.00016 vs. 0.109, p-value 2.30E-141), *FYCO1* rs20040076 (AF:0.00 vs. 0.132, p-value 1.14E-179, *XCR1* rs18111802 (AF:0.00016 vs. 0.0991, p-value 1.15E-128), *OAS3* rs2072134 (AF:0.00097 vs. 0.094, p-value 1.40E-97), and *ABO* rs56390333 (AF: 0.00 vs. 0.0760, p-value 2.47E-103), adding to the current pool of COVID-19 relevant variants; except for *OAS3* rs2072134 which has been associated to metabolic diseases (Heo et al., 2014), these variants have no previous reports. Summarizing, this last analysis showed 287 and 734 potentially protective or deleterious variants on 14 COVID-related genes.

## Discussion

Since 2019, SARS-CoV-2 infection has shown several waves which in some countries have been buffered by vaccination (Zheng et al., 2022). COVID-19 presents an unpredictable course from asymptomatic, mild, or moderate symptoms, to serious complications and 5.5 million deaths. Similar to other viral diseases certain comorbidities are well known to worsen the outcome. COVID-19 patients have an increased risk to be hospitalized if diabetic, hypertensive, or obese. According to the WHO and the Institutional Repository for Information Sharing (IRIS), Mexicans, compared to other countries show an increased COVID-19 risk that ranges from 25-fold for 0.2% of the population to 1.2-fold for 18% of the country’s population (www.iris.paho.org). And although there is no clear protocol to define who is at a specific risk for SARS-CoV-2 infection, defining a genetic predisposition together with clinical and environmental factors could improve diagnosis, treatment, and the development of preventive interventions for high-risk groups potentially using genotype-based diagnosis. Scientific efforts are trying to underpin these genetic and non-genetic factors currently contributing to the collection of data to better depict the disease and to direct personalized care in the years to come.

An interesting review by Z. Yildirim et al. noted that for certain populations higher COVID-19 incidence cannot be explained by behavior, environment, vitamin D levels, socioeconomic status, and cardiovascular risk, but that there is an apparent familial clustering of severity, prompting to genetic variants that could be shared or private for certain ethnic groups (Yildirim et al., 2021). In this study we aimed to assess the allele frequency of variants on 14 COVID-related genes in Mexican Mestizos, and to compare these frequencies between the general population and confirmed COVID-19 patients. At the time of our analysis GWAS analyses have reported 14 genetic loci associated to COVID-19, *ABO, SLC6A20, LZTFL1, CCR9, FYCO1, CXR6, XCR1, CCR2, OAS1-3, TYK2, DPP9, IL10RB/IFNAR2* several of these have been confirmed by other research groups (Anastassopoulou, Gkizarioti, Patrinos, & Tsakris, 2020; Callaway, 2021; Dite et al., 2021b; K Niemi et al., 2021; Velavan et al., 2021), so we focused our investigation on these loci. We identified and observed AF differences for seven previously reported variants, the most statistically relevant for this analysis were variants *TYK2* rs74956615, *IFNAR2* rs2236757, *DPP9* rs2109069 and those on *OAS1-3* (Table 2). These variants have not been yet defined as causal of SARS-CoV-2 infection or severity, but it is known that they play a relevant role in the immune response (Velavan et al., 2021). *IFNAR* as part of interferons receptor conveys antiviral immunity, TYK2, tyrosine kinase 2 is part of the effector cascade of activated interferon receptors, increases susceptibility to microbial infections and has been recently suggested as a drug target (K Niemi et al., 2021; Pairo-Castineira et al., 2020b), *DPP9* codes for a dipeptidyl peptidase from the family of serine proteases similar to DPP4 also known as CD26 that prompts T cell activation. Genetic variants on both genes *DPP4* and *DPP9* (ex. rs13015258 and rs2109069) have been associated with severe COVID-19 (Asgari & Pousaz, 2021; Yildirim et al., 2021). Finally, *OAS1-3* genes are considered “antiviral” as their products trigger viral mRNA degradation and are induced by interferons. Variants on these genes and its classification on affecting the immune system may only be useful to appropriately direct patients infected with SARS-Co-V2, as research has shown that specific variation on immunity genes may represent risk for one disease but protection for another, as it happened for the *CCR5* mutation that protects against HIV but increases the risk for West Nile virus infection (Glass et al., 2005). Inflammation and inflammaging, immunosuppression, and genetically immunodeficient are terms depicting COVID-19 severe cases. For example, inflammaging results from the chronic stimulation of the innate immune system with a high damaging impact during aging, it is a common and conserved mechanism of metabolic diseases in the elderly (Dall’Olio et al., 2013; Franceschi, Garagnani, Parini, Giuliani, & Santoro, 2018). Current research ought to identify and dissect genetic variants that associate with these altered-immune stages of the disease and to better define immunodeficiency in genetic terms which will complement the proper identification of patients at risk for severe COVID-19.

Initial reports on the genetics of severe COVID-19 prompted almost immediately to blood type, suggesting that while type O may offer some protection against SARS-CoV-2 infection, types A and B may be present in patients at risk, but this failed to replicate in several studies (Latz et al., 2020). The Severe COVID-19 group reported *ABO* variants, rs8176747, rs657152, and rs41302905 significantly associated to severe COVID-19 (Zhao et al., 2021), for the two former we did not observed allele frequency differences when comparing groups. Nevertheless, we identified 157 variants on ABO with significant AF differences (p-value <0.01). Among those we observed *ABO* rs657152-A differing significantly between COVID-19 patients and the general population (AF: 0.3632 in COVID vs. 0.2436, p-value=5.25E-05), this variant has been associated with risk of deep-vein thrombosis and pulmonary embolism, conditions present in patients with severe COVID-19 (Aydillo, Esther Babady, & Kamboj, 2020b). Our observations may support current reports addressing the lack of a consistent association between ABO blood types and disease severity, which may warrant delving into a comprehensive consideration of the ABO system and its relation to viral infection, immunogenetics, and disease (Fleeson et al., 2017; Latz et al., 2020; Paul et al., 2021; Zhang, Bastard, Liu, et al., 2020).

We observed a lack of confirmation on gene variants previously reported to play a role in the immunity against COVID-19 and antiviral defense (Zeberg & Pääbo, 2021b; Zhang, Bastard, Bolze, et al., 2020) for *IFNAR* rs2236757, *ABO* rs8176747, and *OAS3* rs2285932, which did not show significant allele frequency differences. *IFNAR* rs2236757 showed an AF:0.456 in COVID-19 patients and 0.439 in the general population. This variant is globally frequent in Asians (AF:0.630) and Caucasians (AF:0.300). Castineira et al reported a 1.3 OR towards increased COVID severity when the variant is present, putatively affecting antiviral defense mechanisms, organ damage, and inflammation. For *ABO* rs8176747, previous reports indicate a higher frequency in affected patients, we also observed a 40% higher AF in COVID patients, but here differences did not reach statistical significance. Intronic Neandertal-derived *OAS1* rs4766664 and *OAS3* rs2285932 identified in the GenOMICC study (Pairo-Castineira et al., 2020b; Zeberg & Pääbo, 2020) did not reach statistical significance in our comparisons. Both SNVs showed a lower AF in COVID-19 patients which are lower than AF values reported in Mexicans from Los Angeles suggesting that its protective trait may not be present in this study population. It is possible that for these three previously COVID-19 associated variants the small size of our COVID group is hindering allele frequency differences, or that haplotypes are not in full LD in this admixed population.

### Additional potentially relevant variants

Despite the absence of replicating results for the three above mentioned variants we observed significant AF differences on over 100 variants including, four on *ABO* p-value<E62-103, *IFNAR2* rs144511572 (p-value<E-40), and *OAS1-3* rs16942374, rs2072174 p-value<E-84, *XCR1* rs181118021, *SLC6A20* rs184655598, *CXCR6* rs113318190, rs200040076, *DPP9* rs118149076 rs72977997, and missense *TYK2* rs35018800 p-value<E-34 previously associated with autoimmunity (Diogo et al., 2015), except for the latter, none of these have been associated to disease risk, and most of them are without biomedical information or scientific citation.

Here, we confirmed the association between COVID-19 and seven genetic variants on *ABO, LZTFL1, OAS1-3*, and *DPP9*. Moreover, we provide a list of over 100 variants on loci that have been consistently associated to COVID-19 and that showed significant allele frequency differences between COVID patients and the general population that have not been previously listed. It is important to highlight that the available clinical information and small sample size of COVID-19 patients limited our mathematical and statistical analyses and was not enough for developing association models, the pandemic impacted laboratory resources and computational capacity therefore our endeavors here were directed for allele frequency (AF) comparisons in 14 specific genes/loci on chromosomes 3, 9, 12, 19, and 21 on which COVID-19 associated variants have been consistently reported (Aydillo et al., 2020b; K Niemi et al., 2021; Pairo-Castineira et al., 2020a).

Mexico is within the top 15 countries with the highest number of COVID cases and 4^th^ in COVID-related deaths, infection and mortality are not consistent or in proportion with population density compared to larger countries. Despite vaccination coverage, the high mortality rate in this admixed population highlights the relevance of investigating all potential factors impacting COVID-19 with the consideration of geographical ancestry. It is possible that some of the genetic variants here identified may contribute to the immunogenetics of SARS-CoV-2 infection in Mexican Mestizos only or may be observed in other populations with shared geographical ancestries. Further studies to confirm these observations and to delve into the ethnic and linkage disequilibrium differences to generate risk models are warranted.

## Conclusion

Ongoing research has identified genetic variants associated with COVID-19 with a diversity of roles including cardiovascular health, hypertension, metabolic diseases, and the immune response. Epigenetic studies report a higher impact of genetic factors compared to epigenetic mechanisms for COVID severity (Yildirim et al., 2021). Despite the identification of some relevant genetic variants in T2D, hypertension, and metabolic diseases, the intersection between these variants and those recently reported associated to COVID-19 is barely present. Research on immunogenetics, genetic ancestry, and the genetics of susceptibility to SARS-CoV-2 infection may open research paths leading to understand the intersection between genetics and metabolic pathways of the immune system, inflammation, metabolic dysfunction, inflammaging, and viral infection. Our observations confirm and complement previous reports on allele frequency differences between COVID-19 patients and a general admixed population. Confirmation and validation of the most relevant variants associated to COVID-19 may lead to the development of models that could assess individual COVID-19 risk potentially supporting personalized prevention and treatment.

## Data Availability

All data produced in the present work are contained in the manuscript

## Acknowledgments

We gratefully acknowledge the support of Cintia K. Guzman-Cruz and all the health care personnel from the Unidad de Servicios Externos e Investigación Clínica (USEIC) and UDIMEB at ENCB-IPN for their technical assistance, patient recruitment, and laboratory testing. Tomas Texis is supported by a Conacyt Graduate Scholarship 2021-2023.

## Author contributions

Study design and clinical sampling: S.S-G, MC. J-M, E. P-B, A.N-P, L.A-C, L. H-T, S.M. P-T; Imputation and Bioinformatic analyses: J.L. C-J, W. G-M, T.T; Bioinformatic and statistical analyses: T.T; Clinical database mining and critical review of the paper: J.D. M-E, M. R-D, S. C-R; Conceptualization and wrote the paper: V. G-C.

## Data availability statement

Supplemental Table 1 lists all variants identified on COVID-19 associated genes with an allele frequency ≥0.01%. Raw imputation data are available from the corresponding author upon reasonable request.

