## Supplemental Table 1 for "COVID-19 relevant genetic variants confirmed in an admixed population"

| SNP | MAFNonCovid | MAF COVID | N COVID | p-value-MAF |
|---|---|---|---|---|
| rs56390333 | 0 | 0.07547 | 101 | 2.47E-103 |
| **rs35494115** | 0.000487 | 0.08019 | 103 | 1.86E-92 |
| **rs371569951** | 0.000487 | 0.06132 | 104 | 8.44E-68 |
| **rs200932155** | 0.000974 | 0.06604 | 106 | 3.07E-62 |
| **rs113318190** | 0.000162 | 0.1085 | 95 | 2.30E-141 |
| **rs72977997** | 0.003085 | 0.09906 | 98 | 9.38E-68 |
| **rs118149076** | 0.000812 | 0.07075 | 104 | 1.23E-71 |
| rs200040076 | 0 | 0.1321 | 95 | 1.14E-179 |
| **rs181118021** | 0.000162 | 0.09906 | 98 | 1.15E-128 |
| **rs16942374** | 0.000162 | 0.06604 | 101 | 2.54E-84 |
| **rs2072134** | 0.000974 | 0.09434 | 98 | 1.40E-97 |
| rs7466899 | 0.000974 | 0.03302 | 106 | 2.84E-24 |
| rs8176739 | 0.005196 | 0.07075 | 103 | 5.53E-28 |
| rs1053878 | 0.03654 | 0.2028 | 98 | 1.95E-32 |
| rs8176716 | 0.007957 | 0.1179 | 99 | 7.28E-49 |
| rs9411372 | 0.00065 | 0.03774 | 104 | 1.81E-34 |
| rs8176705 | 0.004547 | 0.07075 | 101 | 5.50E-31 |
| rs61736301 | 0.000162 | 0.0283 | 105 | 5.14E-34 |
| rs543040 | 0.2335 | 0.5708 | 57 | 2.93E-29 |
| rs613534 | 0.2442 | 0.5708 | 56 | 7.93E-27 |
| rs544873 | 0.2442 | 0.566 | 56 | 4.09E-26 |
| rs545971 | 0.2335 | 0.5519 | 59 | 2.67E-26 |
| rs612169 | 0.2335 | 0.5519 | 59 | 2.67E-26 |
| rs597988 | 0.2332 | 0.5519 | 59 | 2.26E-26 |
| rs597974 | 0.2332 | 0.5519 | 59 | 2.26E-26 |
| rs576123 | 0.2332 | 0.5519 | 59 | 2.26E-26 |
| rs8176663 | 0.2335 | 0.5519 | 59 | 2.67E-26 |
| rs491626 | 0.2332 | 0.5425 | 60 | 5.99E-25 |
| rs492488 | 0.2332 | 0.5425 | 60 | 5.99E-25 |
| rs493246 | 0.2332 | 0.5425 | 60 | 5.99E-25 |
| rs494242 | 0.2439 | 0.5519 | 58 | 4.21E-24 |
| rs495203 | 0.2332 | 0.5425 | 60 | 5.99E-25 |
| rs582118 | 0.2161 | 0.5142 | 65 | 1.99E-24 |
| rs582094 | 0.217 | 0.5425 | 62 | 1.12E-28 |
| rs79006711 | 0.0302 | 0.1981 | 90 | 2.70E-38 |
| rs41289608 | 0.03313 | 0.2264 | 88 | 9.74E-46 |
| rs6775854 | 0.03556 | 0.2028 | 92 | 1.76E-33 |
| rs7648467 | 0.03556 | 0.2028 | 92 | 1.76E-33 |
| rs17078408 | 0.03573 | 0.2028 | 92 | 2.65E-33 |
| rs75192040 | 0.03621 | 0.1792 | 95 | 5.17E-25 |
| rs113305890 | 0.02273 | 0.1321 | 96 | 1.54E-22 |
| rs73539528 | 0.02192 | 0.1321 | 96 | 1.74E-23 |
| rs75894880 | 0.01786 | 0.1321 | 96 | 2.87E-29 |
| rs57034092 | 0.02209 | 0.1321 | 96 | 2.73E-23 |
| rs74840661 | 0.01007 | 0.09434 | 99 | 9.93E-27 |

| | | | | |
|---|---|---|---|---|
| rs78788550 | 0.01056 | 0.09434 | 99 | 1.41E-25 |
| rs16992502 | 0.01185 | 0.09434 | 99 | 7.20E-23 |
| rs79945697 | 0.01056 | 0.09906 | 99 | 4.85E-28 |
| rs75191837 | 0.002273 | 0.07547 | 102 | 7.01E-53 |
| rs114625537 | 0.01056 | 0.09434 | 100 | 1.41E-25 |
| rs60431156 | 0.01559 | 0.1038 | 102 | 7.78E-21 |
| rs564601460 | 0.001949 | 0.04245 | 106 | 4.71E-24 |
| rs146554934 | 0.002111 | 0.04245 | 106 | 7.26E-23 |
| rs114548859 | 0.002111 | 0.04245 | 106 | 7.26E-23 |
| rs141155944 | 0.001949 | 0.04245 | 106 | 4.71E-24 |
| rs150785981 | 0.001949 | 0.04245 | 106 | 4.71E-24 |
| rs140002692 | 0.000812 | 0.03774 | 104 | 1.09E-31 |
| rs77896301 | 0.002111 | 0.04245 | 106 | 7.26E-23 |
| rs75726990 | 0.002111 | 0.04245 | 106 | 7.26E-23 |
| rs145828326 | 0.000812 | 0.03774 | 104 | 1.09E-31 |
| rs181518477 | 0.001949 | 0.04245 | 106 | 4.71E-24 |
| rs140228956 | 0.001949 | 0.04245 | 106 | 4.71E-24 |
| rs189847748 | 0.001949 | 0.04245 | 106 | 4.71E-24 |
| rs141064206 | 0 | 0.02358 | 106 | 1.80E-33 |
| rs141476300 | 0 | 0.02358 | 106 | 1.80E-33 |
| rs7619256 | 0.03264 | 0.1934 | 92 | 2.24E-33 |
| rs3947589 | 0.06772 | 0.2406 | 90 | 2.44E-21 |
| rs144511372 | 0 | 0.0283 | 106 | 7.91E-40 |
| rs61732395 | 0.001462 | 0.03774 | 105 | 7.84E-24 |
| rs184655598 | 0 | 0.03302 | 105 | 3.50E-46 |
| rs35018800 | 0.002761 | 0.06132 | 102 | 1.79E-34 |
| rs55882956 | 0.0203 | 0.1415 | 97 | 2.02E-29 |
| rs569981396 | 0 | 0.00472 | 106 | 7.05E-08 |
| rs10901251 | 0.1431 | 0.3208 | 88 | 9.06E-13 |
| rs34266669 | 0.05278 | 0.1274 | 99 | 2.97E-06 |
| rs199969472 | 0.05278 | 0.1274 | 99 | 2.97E-06 |
| rs35184739 | 0.05261 | 0.1274 | 99 | 2.74E-06 |
| rs7849280 | 0.05294 | 0.1274 | 99 | 3.22E-06 |
| rs9411475 | 0.04449 | 0.1179 | 99 | 6.69E-07 |
| rs13291798 | 0.0354 | 0.1085 | 100 | 4.15E-08 |
| rs34085694 | 0.06073 | 0.1792 | 100 | 5.28E-12 |
| rs4962113 | 0.4209 | 0.3349 | 89 | 1.26E-02 |
| rs111590440 | 0.01104 | 0.0283 | 106 | 2.11E-02 |
| rs73660468 | 0.008282 | 0.04717 | 106 | 1.09E-08 |
| rs7870156 | 0.01023 | 0.04245 | 106 | 1.28E-05 |
| rs62574565 | 0.4638 | 0.5613 | 66 | 5.13E-03 |
| rs58081338 | 0.4729 | 0.6274 | 59 | 9.54E-06 |
| rs62574567 | 0.4729 | 0.6274 | 59 | 9.54E-06 |
| rs60484807 | 0.4688 | 0.3255 | 91 | 3.87E-05 |
| rs12554580 | 0.4688 | 0.3302 | 90 | 6.91E-05 |
| rs12554336 | 0.4726 | 0.6274 | 59 | 9.14E-06 |

| | | | | |
|---|---|---|---|---|
| rs12554339 | 0.4657 | 0.5755 | 66 | 1.65E-03 |
| rs10901253 | 0.4726 | 0.6274 | 59 | 9.14E-06 |
| rs10751502 | 0.4657 | 0.3208 | 92 | 3.13E-05 |
| rs11244052 | 0.4657 | 0.5755 | 66 | 1.65E-03 |
| rs11244053 | 0.4726 | 0.6274 | 59 | 9.14E-06 |
| rs4962114 | 0.4726 | 0.6274 | 59 | 9.14E-06 |
| rs4962115 | 0.4726 | 0.6274 | 59 | 9.14E-06 |
| rs4962116 | 0.4726 | 0.6274 | 59 | 9.14E-06 |
| rs8176757 | 0.4696 | 0.3302 | 90 | 6.27E-05 |
| rs111207633 | 0.00682 | 0.0283 | 106 | 3.76E-04 |
| rs11244054 | 0.3157 | 0.2264 | 99 | 5.84E-03 |
| rs530909128 | 0.000325 | 0.004717 | 106 | 3.75E-03 |
| rs7469795 | 0.4701 | 0.3255 | 91 | 3.30E-05 |
| rs7466519 | 0.00682 | 0.0283 | 106 | 3.76E-04 |
| rs8176748 | 0.4657 | 0.5802 | 66 | 1.03E-03 |
| rs8176745 | 0.4662 | 0.5802 | 66 | 1.08E-03 |
| rs201439325 | 0.000487 | 0.004717 | 106 | 1.56E-02 |
| rs56116432 | 0.00065 | 0.004717 | 106 | 3.76E-02 |
| rs557530257 | 0 | 0.00472 | 106 | 7.05E-08 |
| rs7855255 | 0.06642 | 0.1085 | 102 | 1.66E-02 |
| rs8176735 | 0.04027 | 0.009434 | 106 | 2.30E-02 |
| rs8176733 | 0.06642 | 0.1085 | 102 | 1.66E-02 |
| rs2073823 | 0.06642 | 0.1085 | 102 | 1.66E-02 |
| rs8176730 | 0.06642 | 0.1085 | 102 | 1.66E-02 |
| rs8176729 | 0.04027 | 0.009434 | 106 | 2.30E-02 |
| rs8176725 | 0.0669 | 0.1085 | 102 | 1.83E-02 |
| rs111310794 | 0.06155 | 0.01415 | 106 | 4.27E-03 |
| rs45610939 | 0.009256 | 0.02358 | 105 | 3.67E-02 |
| rs4962040 | 0.2092 | 0.1509 | 99 | 3.98E-02 |
| rs138164693 | 0.008769 | 0.02358 | 105 | 2.68E-02 |
| rs8176710 | 0.0669 | 0.01415 | 106 | 2.21E-03 |
| rs183228393 | 0.008769 | 0.02358 | 105 | 2.68E-02 |
| rs8176702 | 0.2082 | 0.1321 | 100 | 7.02E-03 |
| rs8176698 | 0.00406 | 0.01415 | 106 | 2.90E-02 |
| rs8176696 | 0.00406 | 0.01415 | 106 | 2.90E-02 |
| rs687621 | 0.233 | 0.3019 | 86 | 2.01E-02 |
| rs687289 | 0.2319 | 0.3019 | 86 | 1.80E-02 |
| rs150372066 | 0.00341 | 0.0283 | 106 | 4.13E-08 |
| rs143174772 | 0.009419 | 0.06604 | 103 | 1.75E-14 |
| rs151155628 | 0.009419 | 0.06604 | 103 | 1.75E-14 |
| rs8176694 | 0.2186 | 0.1368 | 104 | 4.43E-03 |
| rs150326069 | 0.000162 | 0.004717 | 106 | 2.33E-04 |
| rs8176692 | 0.01072 | 0.06604 | 103 | 1.15E-12 |
| rs145090216 | 0.009419 | 0.06604 | 103 | 1.75E-14 |
| rs8176691 | 0.2134 | 0.1415 | 101 | 1.17E-02 |
| rs8176687 | 0.06479 | 0.01415 | 106 | 2.87E-03 |

| | | | | |
|---|---|---|---|---|
| rs8176684 | 0.007957 | 0.04717 | 106 | 4.62E-09 |
| rs147279040 | 0.0112 | 0.0566 | 103 | 6.61E-09 |
| rs657152 | 0.2436 | 0.3632 | 82 | 7.25E-05 |
| rs8176682 | 0.2135 | 0.1274 | 102 | 2.48E-03 |
| rs139568229 | 0.01072 | 0.0566 | 103 | 2.33E-09 |
| rs149756392 | 0.01072 | 0.0566 | 103 | 2.33E-09 |
| rs145595975 | 0.01072 | 0.0566 | 103 | 2.33E-09 |
| rs148824570 | 0.01072 | 0.0566 | 103 | 2.33E-09 |
| rs146458069 | 0.01072 | 0.0566 | 103 | 2.33E-09 |
| rs181170522 | 0 | 0.00472 | 106 | 7.05E-08 |
| rs141515001 | 0.01072 | 0.0566 | 103 | 2.33E-09 |
| rs8176679 | 0.06479 | 0.01415 | 106 | 2.87E-03 |
| rs587736740 | 0.01072 | 0.05189 | 103 | 6.92E-08 |
| rs138313692 | 0.000325 | 0.01415 | 106 | 1.58E-12 |
| rs149567216 | 0.01072 | 0.0566 | 103 | 2.33E-09 |
| rs8176676 | 0.000162 | 0.009434 | 106 | 9.50E-10 |
| rs143159728 | 0.01072 | 0.0566 | 103 | 2.33E-09 |
| rs587682147 | 0.01072 | 0.05189 | 103 | 6.92E-08 |
| rs587774481 | 0.01072 | 0.05189 | 103 | 6.92E-08 |
| rs587635767 | 0.01056 | 0.03774 | 104 | 2.56E-04 |
| rs1752339 | 0.4602 | 0.5425 | 66 | 1.82E-02 |
| rs148667000 | 0.000325 | 0.01415 | 106 | 1.58E-12 |
| rs142141716 | 0.01072 | 0.05189 | 103 | 6.92E-08 |
| rs587716454 | 0.01072 | 0.05189 | 103 | 6.92E-08 |
| rs587682443 | 0.001949 | 0.01887 | 106 | 1.31E-06 |
| rs587764387 | 0.001949 | 0.01887 | 106 | 1.31E-06 |
| rs587637585 | 0.001949 | 0.01887 | 106 | 1.31E-06 |
| rs144140881 | 0.01072 | 0.05189 | 103 | 6.92E-08 |
| rs1927315 | 0.4602 | 0.5425 | 66 | 1.82E-02 |
| rs514659 | 0.2335 | 0.3679 | 83 | 6.23E-06 |
| rs644234 | 0.2442 | 0.4292 | 75 | 9.75E-10 |
| rs140796254 | 0.01072 | 0.05189 | 103 | 6.92E-08 |
| rs143309559 | 0.01072 | 0.05189 | 103 | 6.92E-08 |
| rs643434 | 0.2442 | 0.4292 | 75 | 9.75E-10 |
| rs145489959 | 0.000325 | 0.01415 | 106 | 1.58E-12 |
| rs587698906 | 0.000487 | 0.004717 | 106 | 1.56E-02 |
| rs139081859 | 0.000325 | 0.01415 | 106 | 1.58E-12 |
| rs142956930 | 0.006983 | 0.03302 | 106 | 2.40E-05 |
| rs8176668 | 0.2278 | 0.07547 | 104 | 1.59E-07 |
| rs574311 | 0.4615 | 0.6415 | 52 | 2.41E-07 |
| rs587663040 | 0 | 0.00472 | 106 | 7.05E-08 |
| rs488775 | 0.4588 | 0.6415 | 52 | 1.56E-07 |
| rs7036642 | 0.2116 | 0.07547 | 104 | 1.51E-06 |
| rs8176661 | 0.00747 | 0.05189 | 106 | 1.46E-11 |
| rs596141 | 0.4682 | 0.3538 | 83 | 1.02E-03 |
| rs587763606 | 0.000487 | 0.004717 | 106 | 1.56E-02 |

| | | | | |
|---|---|---|---|---|
| rs587707823 | 0.1853 | 0.3962 | 81 | 1.83E-14 |
| rs34357864 | 0.2334 | 0.4245 | 79 | 1.45E-10 |
| rs2769071 | 0.2321 | 0.4292 | 79 | 3.52E-11 |
| rs7041458 | 0.00747 | 0.05189 | 106 | 1.46E-11 |
| rs677355 | 0.2321 | 0.4198 | 80 | 2.81E-10 |
| rs676457 | 0.2317 | 0.4151 | 81 | 7.03E-10 |
| rs79343853 | 0.05083 | 0.01415 | 106 | 1.56E-02 |
| rs527210 | 0.2314 | 0.4009 | 82 | 1.15E-08 |
| rs675201 | 0.4547 | 0.5472 | 67 | 7.88E-03 |
| rs550057 | 0.1652 | 0.283 | 90 | 6.73E-06 |
| rs674302 | 0.2332 | 0.4057 | 81 | 7.04E-09 |
| rs551100 | 0.4562 | 0.5519 | 66 | 5.96E-03 |
| rs663054 | 0.4454 | 0.5425 | 68 | 5.23E-03 |
| rs7046674 | 0.2119 | 0.1226 | 102 | 1.67E-03 |
| rs554833 | 0.2324 | 0.4057 | 81 | 5.75E-09 |
| rs8176649 | 0.209 | 0.1226 | 102 | 2.24E-03 |
| rs660340 | 0.2777 | 0.4575 | 75 | 1.12E-08 |
| rs581107 | 0.2778 | 0.4575 | 75 | 1.16E-08 |
| rs8176647 | 0.06138 | 0.009434 | 106 | 1.69E-03 |
| rs659104 | 0.2775 | 0.4151 | 75 | 1.21E-05 |
| rs647800 | 0.2882 | 0.4151 | 75 | 6.55E-05 |
| rs473533 | 0.2775 | 0.4151 | 75 | 1.21E-05 |
| rs147960974 | 0.000162 | 0.004717 | 106 | 2.33E-04 |
| rs475419 | 0.2766 | 0.4104 | 76 | 2.02E-05 |
| rs476410 | 0.2775 | 0.4151 | 75 | 1.21E-05 |
| rs645982 | 0.2775 | 0.4151 | 75 | 1.21E-05 |
| rs500498 | 0.2775 | 0.4151 | 75 | 1.21E-05 |
| rs145218729 | 0.000162 | 0.004717 | 106 | 2.33E-04 |
| rs8176644 | 0.04953 | 0.01415 | 106 | 1.82E-02 |
| rs505922 | 0.2283 | 0.3302 | 82 | 5.48E-04 |
| rs507666 | 0.1374 | 0.2453 | 88 | 9.04E-06 |
| rs529565 | 0.229 | 0.3349 | 81 | 3.31E-04 |
| rs8176641 | 0.05992 | 0.009434 | 106 | 2.03E-03 |
| rs532436 | 0.1811 | 0.2406 | 90 | 2.76E-02 |
| rs8176639 | 0.05992 | 0.009434 | 106 | 2.03E-03 |
| rs8176760 | 0.000325 | 0.009434 | 106 | 1.93E-07 |
| rs3749461 | 0.02842 | 0.1179 | 95 | 2.02E-13 |
| rs3918362 | 0.02842 | 0.1179 | 95 | 2.02E-13 |
| rs3092963 | 0.2892 | 0.4198 | 81 | 4.04E-05 |
| rs3918365 | 0.02842 | 0.1179 | 95 | 2.02E-13 |
| rs1799865 | 0.2892 | 0.4198 | 81 | 4.04E-05 |
| rs3138042 | 0.2892 | 0.4198 | 81 | 4.04E-05 |
| rs140253702 | 0.000325 | 0.004717 | 106 | 3.75E-03 |
| rs1860264 | 0.2621 | 0.4528 | 77 | 7.20E-10 |
| rs186153536 | 0.000325 | 0.004717 | 106 | 3.75E-03 |
| rs74808020 | 0.0354 | 0.08491 | 102 | 1.75E-04 |
| rs530595037 | 0.000325 | 0.004717 | 106 | 3.75E-03 |

| | | | | |
|---|---|---|---|---|
| rs552329766 | 0.000325 | 0.004717 | 106 | 3.75E-03 |
| rs12631321 | 0.0354 | 0.08491 | 102 | 1.75E-04 |
| rs150667470 | 0.000487 | 0.004717 | 106 | 1.56E-02 |
| rs6441930 | 0.2621 | 0.4528 | 77 | 7.20E-10 |
| rs138421181 | 0 | 0.00472 | 106 | 7.05E-08 |
| rs115004584 | 0.000325 | 0.004717 | 106 | 3.75E-03 |
| rs78414305 | 0.000325 | 0.004717 | 106 | 3.75E-03 |
| rs115826613 | 0 | 0.00472 | 106 | 7.05E-08 |
| rs74739895 | 0.000325 | 0.004717 | 106 | 3.75E-03 |
| rs75018424 | 0.03524 | 0.08491 | 102 | 1.62E-04 |
| rs78044818 | 0.000325 | 0.004717 | 106 | 3.75E-03 |
| rs74337052 | 0 | 0.00472 | 106 | 7.05E-08 |
| rs7636844 | 0.1406 | 0.08491 | 106 | 2.10E-02 |
| rs2286486 | 0.1406 | 0.08019 | 106 | 1.23E-02 |
| rs116350857 | 0 | 0.00472 | 106 | 7.05E-08 |
| rs6441931 | 0.1406 | 0.08491 | 106 | 2.10E-02 |
| rs34338823 | 0.04141 | 0.01415 | 106 | 4.78E-02 |
| rs115227401 | 0.000325 | 0.004717 | 106 | 3.75E-03 |
| rs76478208 | 0.000325 | 0.004717 | 106 | 3.75E-03 |
| rs6783694 | 0.000325 | 0.004717 | 106 | 3.75E-03 |
| rs6441932 | 0.272 | 0.4481 | 77 | 1.82E-08 |
| rs116264461 | 0.000325 | 0.004717 | 106 | 3.75E-03 |
| rs77676459 | 0.000325 | 0.004717 | 106 | 3.75E-03 |
| rs114406614 | 0 | 0.00472 | 106 | 7.05E-08 |
| rs144180015 | 0.000325 | 0.004717 | 106 | 3.75E-03 |
| rs9862611 | 0.2714 | 0.4481 | 77 | 1.58E-08 |
| rs71325091 | 0.04141 | 0.01415 | 106 | 4.78E-02 |
| rs6441933 | 0.272 | 0.4481 | 77 | 1.82E-08 |
| rs141167007 | 0 | 0.00472 | 106 | 7.05E-08 |
| rs57527954 | 0.272 | 0.4481 | 77 | 1.82E-08 |
| rs9834860 | 0.272 | 0.4481 | 77 | 1.82E-08 |
| rs116207980 | 0 | 0.00472 | 106 | 7.05E-08 |
| rs12638201 | 0.03037 | 0.07547 | 102 | 2.37E-04 |
| rs9869542 | 0.1507 | 0.08491 | 106 | 8.08E-03 |
| rs56058226 | 0.3823 | 0.3113 | 88 | 3.64E-02 |
| rs75078036 | 0.02939 | 0.07075 | 102 | 6.06E-04 |
| rs9855205 | 0.05213 | 0.009434 | 106 | 5.34E-03 |
| rs3764864 | 0.07373 | 0.03774 | 106 | 4.70E-02 |
| rs6782814 | 0.08672 | 0.2264 | 92 | 3.89E-12 |
| rs9823523 | 0.05602 | 0.02358 | 106 | 4.15E-02 |
| rs3774642 | 0.02923 | 0.07547 | 102 | 1.26E-04 |
| rs3774641 | 0.1172 | 0.05189 | 106 | 3.37E-03 |
| rs1488371 | 0.2761 | 0.4434 | 79 | 1.01E-07 |
| rs147753307 | 0.001624 | 0.01415 | 106 | 7.07E-05 |
| rs2236938 | 0.1159 | 0.25 | 92 | 3.70E-09 |
| rs17764980 | 0.04157 | 0.01415 | 106 | 4.69E-02 |

| | | | | |
|---|---|---|---|---|
| rs79663567 | 0 | 0.00472 | 106 | 7.05E-08 |
| rs73830609 | 0.002436 | 0.01415 | 106 | 1.58E-03 |
| rs17714101 | 0.04157 | 0.01415 | 106 | 4.69E-02 |
| rs1985356 | 0.1567 | 0.3538 | 85 | 2.29E-14 |
| rs1985463 | 0.207 | 0.3821 | 85 | 9.28E-10 |
| rs59817490 | 0.003897 | 0.01415 | 106 | 2.39E-02 |
| rs7614342 | 0.1565 | 0.3538 | 85 | 2.14E-14 |
| rs79587259 | 0.02907 | 0.07075 | 102 | 5.15E-04 |
| rs61751653 | 0.001786 | 0.009434 | 105 | 1.53E-02 |
| rs74718475 | 0 | 0.00472 | 106 | 7.05E-08 |
| rs73830610 | 0.003897 | 0.01415 | 106 | 2.39E-02 |
| rs17714228 | 0.04157 | 0.01415 | 106 | 4.69E-02 |
| rs875891 | 0.1554 | 0.3538 | 85 | 1.29E-14 |
| rs875890 | 0.2069 | 0.3821 | 85 | 8.86E-10 |
| rs62245098 | 0.1565 | 0.3585 | 84 | 5.20E-15 |
| rs28595837 | 0.02225 | 0.05189 | 102 | 4.85E-03 |
| rs9881306 | 0.05213 | 0.009434 | 106 | 5.34E-03 |
| rs28501674 | 0.1552 | 0.3538 | 85 | 1.20E-14 |
| rs61565453 | 0.1565 | 0.3585 | 84 | 5.20E-15 |
| rs71325092 | 0.04157 | 0.01415 | 106 | 4.69E-02 |
| rs79179998 | 0.05213 | 0.009434 | 106 | 5.34E-03 |
| rs57295094 | 0.1549 | 0.3538 | 85 | 1.03E-14 |
| rs6791859 | 0.1609 | 0.3443 | 86 | 1.93E-12 |
| rs4683147 | 0.3966 | 0.566 | 65 | 7.49E-07 |
| rs74945326 | 0.04108 | 0.009434 | 106 | 2.08E-02 |
| rs62245101 | 0.1146 | 0.2311 | 95 | 2.48E-07 |
| rs62245102 | 0.1593 | 0.3396 | 87 | 3.67E-12 |
| rs7638236 | 0.019 | 0.04245 | 105 | 1.59E-02 |
| rs4535265 | 0.4108 | 0.5094 | 71 | 4.17E-03 |
| rs9869848 | 0.1111 | 0.1887 | 93 | 4.65E-04 |
| rs9875616 | 0.109 | 0.1887 | 93 | 2.90E-04 |
| rs13091979 | 0.4105 | 0.5094 | 71 | 4.04E-03 |
| rs13091821 | 0.4105 | 0.5094 | 71 | 4.04E-03 |
| rs17078449 | 0.1471 | 0.08019 | 105 | 6.48E-03 |
| rs936938 | 0.4074 | 0.5 | 72 | 7.07E-03 |
| rs2171531 | 0.04579 | 0.009434 | 106 | 1.17E-02 |
| rs17078454 | 0.1466 | 0.08019 | 105 | 6.81E-03 |
| rs57204020 | 0.1465 | 0.08019 | 105 | 6.92E-03 |
| rs7634640 | 0.07616 | 0.1651 | 94 | 2.40E-06 |
| rs55920693 | 0.04579 | 0.009434 | 106 | 1.17E-02 |
| rs3774640 | 0.1465 | 0.08019 | 105 | 6.92E-03 |
| rs3774639 | 0.4105 | 0.5094 | 71 | 4.04E-03 |
| rs3774638 | 0.1465 | 0.08019 | 105 | 6.92E-03 |
| rs7627147 | 0.01851 | 0.04245 | 105 | 1.27E-02 |
| rs2234350 | 0.1259 | 0.1887 | 93 | 7.09E-03 |
| rs2234351 | 0.1462 | 0.08019 | 105 | 7.15E-03 |

| | | | | |
|---|---|---|---|---|
| rs191811571 | 0.000162 | 0.004717 | 106 | 2.33E-04 |
| rs3774635 | 0.4074 | 0.5 | 72 | 7.07E-03 |
| rs936939 | 0.1465 | 0.08019 | 105 | 6.92E-03 |
| rs143980719 | 0.001949 | 0.01415 | 106 | 3.13E-04 |
| rs2234355 | 0.02533 | 0.06132 | 105 | 1.35E-03 |
| rs17078464 | 0.01835 | 0.04245 | 105 | 1.18E-02 |
| rs2054866 | 0.4074 | 0.5 | 72 | 7.07E-03 |
| rs58442576 | 0.01851 | 0.04245 | 105 | 1.27E-02 |
| rs4683156 | 0.4105 | 0.5094 | 71 | 4.04E-03 |
| rs56239523 | 0.01851 | 0.04245 | 105 | 1.27E-02 |
| rs58050797 | 0.01835 | 0.04245 | 105 | 1.18E-02 |
| rs1994488 | 0.4074 | 0.5 | 72 | 7.07E-03 |
| rs35501575 | 0.04596 | 0.009434 | 106 | 1.14E-02 |
| rs1552489 | 0.4074 | 0.5 | 72 | 7.07E-03 |
| rs8107316 | 0.2243 | 0.3066 | 88 | 4.89E-03 |
| rs72977966 | 0 | 0.00472 | 106 | 7.05E-08 |
| rs8105807 | 0.3037 | 0.3679 | 84 | 4.59E-02 |
| rs10421782 | 0.3027 | 0.3679 | 84 | 4.25E-02 |
| rs2885734 | 0.3037 | 0.3679 | 84 | 4.59E-02 |
| rs114346470 | 0.000162 | 0.004717 | 106 | 2.33E-04 |
| rs34136596 | 0.00065 | 0.004717 | 106 | 3.76E-02 |
| rs113439332 | 0.0328 | 0.1321 | 96 | 2.58E-14 |
| rs113094386 | 0.005684 | 0.01887 | 105 | 1.55E-02 |
| rs2277735 | 0.2947 | 0.3821 | 82 | 6.25E-03 |
| rs60199752 | 0.2241 | 0.3066 | 87 | 4.80E-03 |
| rs35408525 | 0.00065 | 0.004717 | 106 | 3.76E-02 |
| rs11671605 | 0.2241 | 0.3066 | 87 | 4.80E-03 |
| rs4401152 | 0.2244 | 0.3019 | 88 | 8.11E-03 |
| rs72977989 | 0.2241 | 0.3019 | 88 | 7.82E-03 |
| rs139518006 | 0.000162 | 0.004717 | 106 | 2.33E-04 |
| rs11670257 | 0.3514 | 0.5 | 75 | 8.93E-06 |
| rs2277733 | 0.3365 | 0.25 | 100 | 8.65E-03 |
| rs732631 | 0.2756 | 0.3396 | 89 | 4.06E-02 |
| rs2006231 | 0.4292 | 0.3443 | 97 | 1.40E-02 |
| rs2109069 | 0.2462 | 0.3208 | 89 | 1.35E-02 |
| rs2109070 | 0.2446 | 0.3208 | 89 | 1.14E-02 |
| rs3833278 | 0.3909 | 0.3208 | 88 | 3.94E-02 |
| rs1467941 | 0.3988 | 0.3255 | 88 | 3.18E-02 |
| rs1467942 | 0.404 | 0.3349 | 88 | 4.35E-02 |
| rs56945978 | 0.4256 | 0.3443 | 97 | 1.85E-02 |
| rs758510 | 0.3474 | 0.2783 | 91 | 3.76E-02 |
| rs2277732 | 0.1504 | 0.2925 | 89 | 1.92E-08 |
| rs10420007 | 0.3953 | 0.3208 | 88 | 2.90E-02 |
| rs10420225 | 0.4292 | 0.3443 | 97 | 1.40E-02 |
| rs530191556 | 0.3964 | 0.3255 | 88 | 3.77E-02 |
| rs4683148 | 0.4112 | 0.5377 | 70 | 2.36E-04 |

| | | | | |
|---|---|---|---|---|
| rs2171530 | 0.2196 | 0.316 | 92 | 9.01E-04 |
| rs9831315 | 0.4055 | 0.5283 | 72 | 3.50E-04 |
| rs560966886 | 0.000162 | 0.004717 | 106 | 2.33E-04 |
| rs72622922 | 0.000325 | 0.01415 | 106 | 1.58E-12 |
| rs185588322 | 0.001624 | 0.01415 | 106 | 7.07E-05 |
| rs7129 | 0.4058 | 0.5283 | 72 | 3.63E-04 |
| rs1994492 | 0.04222 | 0.009434 | 106 | 1.81E-02 |
| rs1994493 | 0.04222 | 0.009434 | 106 | 1.81E-02 |
| rs3796373 | 0.2212 | 0.3208 | 91 | 6.34E-04 |
| rs3733103 | 0.4112 | 0.533 | 71 | 4.00E-04 |
| rs75928798 | 0.04206 | 0.009434 | 106 | 1.85E-02 |
| rs2291470 | 0.4057 | 0.5283 | 72 | 3.56E-04 |
| rs11130078 | 0.2204 | 0.3349 | 84 | 8.43E-05 |
| rs148655132 | 0.001624 | 0.01415 | 106 | 7.07E-05 |
| rs11707672 | 0.0112 | 0.04245 | 104 | 4.75E-05 |
| rs6800954 | 0.1502 | 0.09906 | 104 | 3.95E-02 |
| rs140100519 | 0.001624 | 0.01415 | 106 | 7.07E-05 |
| rs2248228 | 0.4094 | 0.5094 | 71 | 3.62E-03 |
| rs60237998 | 0.1512 | 0.08962 | 104 | 1.34E-02 |
| rs60110056 | 0.01543 | 0.04245 | 105 | 2.27E-03 |
| rs58491168 | 0.019 | 0.04245 | 105 | 1.59E-02 |
| rs4683149 | 0.4052 | 0.5047 | 71 | 3.74E-03 |
| rs2373087 | 0.04417 | 0.009434 | 106 | 1.43E-02 |
| rs7631809 | 0.1504 | 0.08962 | 104 | 1.44E-02 |
| rs2373088 | 0.06414 | 0.1557 | 94 | 1.67E-07 |
| rs62245106 | 0.113 | 0.0566 | 105 | 1.02E-02 |
| rs12638598 | 0.207 | 0.1132 | 103 | 8.55E-04 |
| rs35831747 | 0.04417 | 0.009434 | 106 | 1.43E-02 |
| rs77709797 | 0.1166 | 0.0566 | 105 | 7.01E-03 |
| rs1532070 | 0.06431 | 0.1557 | 94 | 1.81E-07 |
| rs6778225 | 0.1484 | 0.08491 | 105 | 1.01E-02 |
| rs13099120 | 0.04417 | 0.009434 | 106 | 1.43E-02 |
| rs6778324 | 0.1483 | 0.08491 | 105 | 1.02E-02 |
| rs60779624 | 0.1484 | 0.08491 | 105 | 1.01E-02 |
| rs190039371 | 0.1166 | 0.0566 | 105 | 7.01E-03 |
| rs41289614 | 0.03459 | 0.009434 | 106 | 4.61E-02 |
| rs35477280 | 0.04417 | 0.009434 | 106 | 1.43E-02 |
| rs4683152 | 0.4094 | 0.5094 | 71 | 3.62E-03 |
| rs13066516 | 0.04417 | 0.009434 | 106 | 1.43E-02 |
| rs200582580 | 0.04401 | 0.009434 | 106 | 1.45E-02 |
| rs1072755 | 0.4055 | 0.5 | 72 | 5.92E-03 |
| rs77902290 | 0.04417 | 0.009434 | 106 | 1.43E-02 |
| rs532777636 | 0.04579 | 0.009434 | 106 | 1.17E-02 |
| rs62242787 | 0.1543 | 0.07547 | 105 | 1.66E-03 |
| rs17078471 | 0.002111 | 0.009434 | 106 | 3.06E-02 |
| rs35257780 | 0.04628 | 0.009434 | 106 | 1.10E-02 |
| rs34000569 | 0.04628 | 0.009434 | 106 | 1.10E-02 |

| | | | | |
|---|---|---|---|---|
| rs34324101 | 0.04628 | 0.009434 | 106 | 1.10E-02 |
| rs13069079 | 0.04628 | 0.009434 | 106 | 1.10E-02 |
| rs34849862 | 0.04628 | 0.009434 | 106 | 1.10E-02 |
| rs62242788 | 0.1263 | 0.0566 | 105 | 2.46E-03 |
| rs35209528 | 0.04628 | 0.009434 | 106 | 1.10E-02 |
| rs75918913 | 0.03995 | 0.009434 | 106 | 2.39E-02 |
| rs57036895 | 0.01543 | 0.03774 | 106 | 1.13E-02 |
| rs13079869 | 0.04677 | 0.009434 | 106 | 1.04E-02 |
| rs33910087 | 0.04693 | 0.009434 | 106 | 1.01E-02 |
| rs3796376 | 0.1354 | 0.0566 | 105 | 8.83E-04 |
| rs17215008 | 0.04644 | 0.009434 | 106 | 1.08E-02 |
| rs71325098 | 0.04644 | 0.009434 | 106 | 1.08E-02 |
| rs71325100 | 0.04644 | 0.009434 | 106 | 1.08E-02 |
| rs41289622 | 0.04644 | 0.009434 | 106 | 1.08E-02 |
| rs11130081 | 0.002111 | 0.009434 | 106 | 3.06E-02 |
| rs12639598 | 0.405 | 0.5047 | 73 | 3.68E-03 |
| rs144873702 | 0.000812 | 0.01415 | 106 | 6.97E-08 |
| rs751552 | 0.4053 | 0.5142 | 72 | 1.53E-03 |
| rs751553 | 0.4053 | 0.5142 | 72 | 1.53E-03 |
| rs904634 | 0.08379 | 0.1604 | 96 | 9.48E-05 |
| rs13066062 | 0.04644 | 0.009434 | 106 | 1.08E-02 |
| rs6781668 | 0.1351 | 0.0566 | 105 | 9.17E-04 |
| rs9844821 | 0.07161 | 0.1509 | 101 | 1.48E-05 |
| rs57228214 | 0.02192 | 0.05189 | 106 | 4.15E-03 |
| rs9849771 | 0.09337 | 0.1604 | 96 | 1.11E-03 |
| rs9849818 | 0.08396 | 0.1604 | 96 | 9.94E-05 |
| rs4682801 | 0.1291 | 0.2547 | 88 | 1.22E-07 |
| rs35537559 | 0.1971 | 0.09434 | 105 | 1.96E-04 |
| rs59767267 | 0.1314 | 0.0566 | 105 | 1.40E-03 |
| rs3821883 | 0.1346 | 0.0566 | 105 | 9.69E-04 |
| rs36122610 | 0.04628 | 0.009434 | 106 | 1.10E-02 |
| rs138017682 | 0.00065 | 0.009434 | 106 | 4.14E-05 |
| rs34442130 | 0.04628 | 0.009434 | 106 | 1.10E-02 |
| rs931704 | 0.4039 | 0.4764 | 77 | 3.45E-02 |
| rs4683159 | 0.203 | 0.316 | 87 | 6.43E-05 |
| rs13075758 | 0.04628 | 0.009434 | 106 | 1.10E-02 |
| rs11130082 | 0.2035 | 0.3208 | 86 | 3.46E-05 |
| rs6802312 | 0.1159 | 0.2547 | 88 | 1.05E-09 |
| rs56972507 | 0.02144 | 0.04717 | 106 | 1.26E-02 |
| rs3733097 | 0.4058 | 0.5189 | 72 | 9.98E-04 |
| rs17078495 | 0.4058 | 0.5189 | 72 | 9.98E-04 |
| rs13086858 | 0.2035 | 0.3208 | 86 | 3.46E-05 |
| rs139886125 | 0.000812 | 0.01415 | 106 | 6.97E-08 |
| rs737452 | 0.4058 | 0.5189 | 72 | 9.98E-04 |
| rs730983 | 0.4039 | 0.4906 | 75 | 1.15E-02 |
| rs59226168 | 0.1898 | 0.1038 | 105 | 1.58E-03 |
| rs67044455 | 0.4058 | 0.533 | 70 | 2.14E-04 |

| | | | | |
|---|---|---|---|---|
| rs62242796 | 0.1898 | 0.1038 | 105 | 1.58E-03 |
| rs7653682 | 0.1159 | 0.2877 | 86 | 5.35E-14 |
| rs72891712 | 0.02257 | 0.07547 | 104 | 8.88E-07 |
| rs1873000 | 0.203 | 0.3538 | 83 | 1.05E-07 |
| rs7628447 | 0.2035 | 0.3538 | 83 | 1.18E-07 |
| rs12635657 | 0.4057 | 0.5283 | 70 | 3.56E-04 |
| rs2133660 | 0.2371 | 0.3774 | 81 | 2.73E-06 |
| rs146216196 | 0.001786 | 0.01887 | 106 | 4.53E-07 |
| rs139044419 | 0.001786 | 0.01887 | 106 | 4.53E-07 |
| rs1846615 | 0.2371 | 0.3774 | 81 | 2.73E-06 |
| rs1873001 | 0.2371 | 0.3774 | 81 | 2.73E-06 |
| rs142154067 | 0.001786 | 0.01887 | 106 | 4.53E-07 |
| rs146285477 | 0.04628 | 0.009434 | 106 | 1.10E-02 |
| rs182171107 | 0.00065 | 0.009434 | 106 | 4.14E-05 |
| rs1500003 | 0.2371 | 0.3774 | 81 | 2.73E-06 |
| rs79067698 | 0.001786 | 0.01887 | 106 | 4.53E-07 |
| rs1392288 | 0.1673 | 0.3349 | 81 | 2.19E-10 |
| rs17330872 | 0.04628 | 0.009434 | 106 | 1.10E-02 |
| rs188114046 | 0.001786 | 0.01887 | 106 | 4.53E-07 |
| rs142571332 | 0.04579 | 0.009434 | 106 | 1.17E-02 |
| rs71327003 | 0.04612 | 0.009434 | 106 | 1.12E-02 |
| rs188401375 | 0.00065 | 0.004717 | 106 | 3.76E-02 |
| rs17860118 | 0.01754 | 0.08962 | 100 | 1.55E-13 |
| rs566208153 | 0.02907 | 0.0566 | 103 | 2.08E-02 |
| rs13052526 | 0.01608 | 0.04717 | 103 | 5.98E-04 |
| rs574424283 | 0.4208 | 0.3491 | 83 | 3.75E-02 |
| rs2248420 | 0.4227 | 0.3443 | 83 | 2.30E-02 |
| rs189110596 | 0.002273 | 0.01415 | 105 | 9.81E-04 |
| rs7509997 | 0.02452 | 0.0566 | 103 | 3.63E-03 |
| rs576036480 | 0.000325 | 0.004717 | 106 | 3.75E-03 |
| rs17860142 | 0.4229 | 0.3443 | 83 | 2.28E-02 |
| rs8130779 | 0.02631 | 0.0566 | 103 | 7.79E-03 |
| rs9974669 | 0.02631 | 0.0566 | 103 | 7.79E-03 |
| rs570824030 | 0.02663 | 0.0566 | 103 | 8.85E-03 |
| rs3153 | 0.4227 | 0.3443 | 83 | 2.30E-02 |
| rs1187 | 0.000487 | 0.004717 | 106 | 1.56E-02 |
| rs12482014 | 0.4229 | 0.3443 | 83 | 2.28E-02 |
| rs17860160 | 0.01608 | 0.04717 | 103 | 5.98E-04 |
| rs12482193 | 0.4285 | 0.3491 | 83 | 2.14E-02 |
| rs17860165 | 0.4229 | 0.3443 | 83 | 2.28E-02 |
| rs117489085 | 0.000325 | 0.004717 | 106 | 3.75E-03 |
| rs545411453 | 0.000325 | 0.004717 | 106 | 3.75E-03 |
| rs8128785 | 0.03215 | 0.0566 | 103 | 5.00E-02 |
| rs2229207 | 0.01803 | 0.06132 | 105 | 7.25E-06 |
| rs1051393 | 0.4329 | 0.316 | 88 | 7.22E-04 |
| rs62226154 | 0.02436 | 0.04717 | 105 | 3.69E-02 |
| rs62226157 | 0.4112 | 0.3396 | 89 | 3.72E-02 |

| | | | | |
|---|---|---|---|---|
| rs144683777 | 0.000812 | 0.01887 | 104 | 5.91E-12 |
| rs10211925 | 0.0229 | 0.04717 | 105 | 2.24E-02 |
| rs17860183 | 0.1669 | 0.04245 | 104 | 1.38E-06 |
| rs2834159 | 0.02079 | 0.04245 | 105 | 3.25E-02 |
| rs12053666 | 0.4104 | 0.2925 | 88 | 5.89E-04 |
| rs2834160 | 0.01689 | 0.04245 | 105 | 5.56E-03 |
| rs2834162 | 0.01786 | 0.04245 | 105 | 9.32E-03 |
| rs17860211 | 0.01786 | 0.04245 | 105 | 9.32E-03 |
| rs2073362 | 0.01689 | 0.06132 | 105 | 2.17E-06 |
| rs17860220 | 0.01673 | 0.06132 | 105 | 1.80E-06 |
| rs147496374 | 0.00341 | 0.02358 | 106 | 5.90E-06 |
| rs4817555 | 0.04027 | 0.1226 | 102 | 6.17E-09 |
| rs59738559 | 0.04108 | 0.1226 | 102 | 1.16E-08 |
| rs2834166 | 0.3357 | 0.2547 | 95 | 1.39E-02 |
| rs11911133 | 0.4732 | 0.3774 | 82 | 5.97E-03 |
| rs181597872 | 0.000325 | 0.004717 | 106 | 3.75E-03 |
| rs2300371 | 0.4048 | 0.2925 | 88 | 1.03E-03 |
| rs1058961 | 0.2408 | 0.4481 | 84 | 6.24E-12 |
| rs35624553 | 0.03638 | 0.009434 | 106 | 3.71E-02 |
| rs139540257 | 0.007145 | 0.02358 | 105 | 7.07E-03 |
| rs2064061 | 0.3459 | 0.5189 | 75 | 2.15E-07 |
| rs67959919 | 0.03638 | 0.009434 | 106 | 3.71E-02 |
| rs148112944 | 0 | 0.00472 | 106 | 7.05E-08 |
| rs11385942 | 0.03832 | 0.009434 | 106 | 2.92E-02 |
| rs11130077 | 0.2311 | 0.3019 | 89 | 1.66E-02 |
| rs6796097 | 0.001299 | 0.009434 | 106 | 3.27E-03 |
| rs190577088 | 0.000487 | 0.004717 | 106 | 1.56E-02 |
| rs35508621 | 0.03459 | 0.004717 | 106 | 1.76E-02 |
| rs758389 | 0.000974 | 0.009434 | 106 | 6.27E-04 |
| rs34288077 | 0.03443 | 0.004717 | 106 | 1.80E-02 |
| rs141146648 | 0.0151 | 0.0566 | 103 | 3.07E-06 |
| rs150719194 | 0.000162 | 0.004717 | 106 | 2.33E-04 |
| rs35081325 | 0.03443 | 0.009434 | 106 | 4.71E-02 |
| rs35731912 | 0.03459 | 0.009434 | 106 | 4.61E-02 |
| rs17078371 | 0.001462 | 0.009434 | 106 | 5.98E-03 |
| rs112101762 | 0.001137 | 0.009434 | 106 | 1.57E-03 |
| rs113181700 | 0.001137 | 0.009434 | 106 | 1.57E-03 |
| rs4683146 | 0.3344 | 0.434 | 83 | 2.57E-03 |
| rs34326463 | 0.03426 | 0.004717 | 106 | 1.84E-02 |
| rs73064425 | 0.03475 | 0.009434 | 106 | 4.52E-02 |
| rs138727962 | 0.000162 | 0.004717 | 106 | 2.33E-04 |
| rs184214751 | 0.000974 | 0.009434 | 106 | 6.27E-04 |
| rs71325089 | 0.02907 | 0.004717 | 106 | 3.54E-02 |
| rs61185149 | 0.008931 | 0.02358 | 106 | 2.99E-02 |
| rs13081482 | 0.03491 | 0.009434 | 106 | 4.43E-02 |
| rs75826707 | 0.003248 | 0.01415 | 105 | 9.26E-03 |

| | | | | |
|---|---|---|---|---|
| rs9871972 | 0.3407 | 0.5 | 73 | 1.63E-06 |
| rs140227473 | 0.001624 | 0.009434 | 106 | 9.92E-03 |
| rs75507082 | 0.02907 | 0 | 106 | 1.18E-02 |
| rs79657519 | 0.001624 | 0.009434 | 106 | 9.92E-03 |
| rs148060386 | 0.000162 | 0.004717 | 106 | 2.33E-04 |
| rs2191031 | 0.1161 | 0.0566 | 105 | 7.38E-03 |
| rs28607988 | 0.04498 | 0.01415 | 106 | 3.13E-02 |
| rs77009309 | 0.02972 | 0.1132 | 100 | 1.58E-11 |
| rs12639224 | 0.138 | 0.2075 | 95 | 4.17E-03 |
| rs12639252 | 0.000487 | 0.004717 | 106 | 1.56E-02 |
| rs2373086 | 0.02842 | 0.1226 | 99 | 1.20E-14 |
| rs112212287 | 0.01624 | 0.06132 | 103 | 1.01E-06 |
| rs7614687 | 0.01072 | 0.0566 | 104 | 2.33E-09 |
| rs57319220 | 0.0544 | 0.02358 | 106 | 4.97E-02 |
| rs34518147 | 0.04125 | 0.01415 | 106 | 4.87E-02 |
| rs9852270 | 0.2621 | 0.4528 | 77 | 7.20E-10 |
| rs560506883 | 0.000162 | 0.004717 | 106 | 2.33E-04 |
| rs77000397 | 0 | 0.00472 | 106 | 7.05E-08 |
| rs114742054 | 0 | 0.00472 | 106 | 7.05E-08 |
| rs12633699 | 0.05505 | 0.01415 | 106 | 9.39E-03 |
| rs34847985 | 0.2131 | 0.3726 | 86 | 3.20E-08 |
| rs34549672 | 0.2811 | 0.4434 | 79 | 2.76E-07 |
| rs75442536 | 0.02273 | 0.0566 | 103 | 1.49E-03 |
| rs59451221 | 0.06041 | 0.1745 | 95 | 2.65E-11 |
| rs9836836 | 0.1341 | 0.0566 | 106 | 1.02E-03 |
| rs35280891 | 0.04157 | 0.009434 | 106 | 1.96E-02 |
| rs181980885 | 0.001624 | 0.01415 | 106 | 7.07E-05 |
| rs186530132 | 0.001624 | 0.01415 | 106 | 7.07E-05 |
| rs112707117 | 0.001624 | 0.01415 | 106 | 7.07E-05 |
| rs34068335 | 0.04206 | 0.009434 | 106 | 1.85E-02 |
| rs558702333 | 0.00065 | 0.004717 | 106 | 3.76E-02 |
| rs35334665 | 0.04726 | 0.009434 | 106 | 9.75E-03 |
| rs115932896 | 0.001299 | 0.01415 | 106 | 9.37E-06 |
| rs71327009 | 0.04726 | 0.009434 | 106 | 9.75E-03 |
| rs189659509 | 0.001299 | 0.009434 | 106 | 3.27E-03 |
| rs35161099 | 0.04726 | 0.009434 | 106 | 9.75E-03 |
| rs12054287 | 0.1323 | 0.05189 | 105 | 6.06E-04 |
| rs60080146 | 0.0177 | 0.03774 | 106 | 3.27E-02 |
| rs59676542 | 0.01819 | 0.03774 | 106 | 3.96E-02 |
| rs58035200 | 0.1958 | 0.1085 | 105 | 1.53E-03 |
| rs7623460 | 0.2658 | 0.1934 | 102 | 1.86E-02 |
| rs71327010 | 0.04726 | 0.009434 | 106 | 9.75E-03 |
| rs7623476 | 0.1957 | 0.1085 | 105 | 1.56E-03 |
| rs7956880 | 0.1728 | 0.08962 | 104 | 1.53E-03 |
| rs10744785 | 0.2512 | 0.1557 | 101 | 1.54E-03 |
| rs4766662 | 0.2407 | 0.1415 | 100 | 8.49E-04 |

| | | | | |
|---|---|---|---|---|
| rs2240190 | 0.03491 | 0.1274 | 98 | 4.43E-12 |
| rs34137742 | 0.07616 | 0.1226 | 103 | 1.29E-02 |
| rs45542343 | 0.03215 | 0.1038 | 102 | 1.97E-08 |
| rs75344264 | 0.1082 | 0.184 | 105 | 5.41E-04 |
| rs2057778 | 0.1772 | 0.1179 | 105 | 2.57E-02 |
| rs2285934 | 0.2459 | 0.1509 | 103 | 1.53E-03 |
| rs4767023 | 0.2621 | 0.1604 | 102 | 8.83E-04 |
| rs45495393 | 0.07567 | 0.1274 | 101 | 5.62E-03 |
| rs56006713 | 0.000974 | 0.009434 | 106 | 6.27E-04 |
| rs12423440 | 0.0492 | 0.08491 | 103 | 1.95E-02 |
| **rs10774671** | **0.275** | **0.184** | **101** | **3.48E-03** |
| rs1131476 | 0.1806 | 0.1132 | 105 | 1.18E-02 |
| **rs2660** | **0.181** | **0.11** | **105** | **7.00E-03** |
| rs7135577 | 0.1806 | 0.1226 | 105 | 3.04E-02 |
| rs4767024 | 0.1801 | 0.1226 | 105 | 3.16E-02 |
| rs4767025 | 0.1801 | 0.1226 | 105 | 3.16E-02 |
| rs4767026 | 0.1807 | 0.1226 | 105 | 3.00E-02 |
| rs4767027 | 0.1801 | 0.1226 | 105 | 3.16E-02 |
| rs4767028 | 0.1806 | 0.1226 | 105 | 3.04E-02 |
| rs4767029 | 0.1801 | 0.1226 | 105 | 3.16E-02 |
| rs75628555 | 0.04758 | 0.08491 | 103 | 1.32E-02 |
| rs4767030 | 0.1806 | 0.1226 | 105 | 3.04E-02 |
| rs10850092 | 0.1801 | 0.1226 | 105 | 3.16E-02 |
| rs6489864 | 0.1801 | 0.1226 | 105 | 3.16E-02 |
| rs6489865 | 0.1801 | 0.1226 | 105 | 3.16E-02 |
| rs10850093 | 0.1801 | 0.1226 | 105 | 3.16E-02 |
| rs10850094 | 0.1801 | 0.1226 | 105 | 3.16E-02 |
| rs10850095 | 0.1801 | 0.1226 | 105 | 3.16E-02 |
| rs10774672 | 0.1807 | 0.1226 | 105 | 3.00E-02 |
| rs10850096 | 0.1801 | 0.1226 | 105 | 3.16E-02 |
| rs10850097 | 0.1981 | 0.1415 | 104 | 4.13E-02 |
| rs10774673 | 0.1801 | 0.1226 | 105 | 3.16E-02 |
| rs10774674 | 0.1801 | 0.1226 | 105 | 3.16E-02 |
| rs3803057 | 0.1312 | 0.06132 | 105 | 2.83E-03 |
| rs1298962 | 0.1702 | 0.09906 | 105 | 6.43E-03 |
| rs1298301 | 0.1632 | 0.2453 | 100 | 1.59E-03 |
| rs1293774 | 0.1702 | 0.09906 | 105 | 6.43E-03 |
| rs117666908 | 0.03361 | 0.06604 | 103 | 1.12E-02 |
| rs1293773 | 0.1629 | 0.2406 | 101 | 2.76E-03 |
| rs1293772 | 0.1819 | 0.08019 | 105 | 1.44E-04 |
| rs531734809 | 0 | 0.00472 | 106 | 7.05E-08 |
| rs1293771 | 0.1819 | 0.08019 | 105 | 1.44E-04 |
| rs1293770 | 0.163 | 0.2358 | 101 | 5.03E-03 |
| rs12301619 | 0.003735 | 0.01415 | 105 | 1.93E-02 |
| rs1293768 | 0.1798 | 0.08019 | 105 | 1.84E-04 |

| | | | | |
|---|---|---|---|---|
| rs150169230 | 0.000162 | 0.004717 | 106 | 2.33E-04 |
| rs1293767 | 0.1259 | 0.06132 | 105 | 5.02E-03 |
| rs1293766 | 0.1575 | 0.2311 | 100 | 4.04E-03 |
| rs1293765 | 0.1645 | 0.07547 | 105 | 5.34E-04 |
| rs1293764 | 0.1639 | 0.07547 | 105 | 5.75E-04 |
| rs1293763 | 0.1523 | 0.2217 | 100 | 6.02E-03 |
| rs11066464 | 0.2085 | 0.2736 | 90 | 2.23E-02 |
| rs757402 | 0.1366 | 0.0566 | 105 | 7.75E-04 |
| rs200629605 | 0.02744 | 0.06132 | 103 | 3.60E-03 |
| rs7975318 | 0.1682 | 0.2311 | 94 | 1.66E-02 |
| rs757401 | 0.1366 | 0.0566 | 105 | 7.75E-04 |
| rs557969944 | 0 | 0.00472 | 106 | 7.05E-08 |
| rs5800979 | 0.3746 | 0.5047 | 70 | 1.24E-04 |
| rs34886194 | 0.2075 | 0.2689 | 90 | 3.10E-02 |
| rs1293762 | 0.1845 | 0.08962 | 102 | 4.24E-04 |
| rs1293760 | 0.1846 | 0.08962 | 102 | 4.17E-04 |
| rs1293759 | 0.1177 | 0.04245 | 105 | 7.37E-04 |
| rs1293758 | 0.2949 | 0.1038 | 102 | 1.55E-09 |
| rs1298961 | 0.3303 | 0.1179 | 102 | 8.00E-11 |
| rs1635142 | 0.1189 | 0.04245 | 105 | 6.45E-04 |
| rs1293757 | 0.1189 | 0.04245 | 105 | 6.45E-04 |
| rs1293756 | 0.1189 | 0.04245 | 105 | 6.45E-04 |
| rs1293754 | 0.1192 | 0.04245 | 105 | 6.20E-04 |
| rs1293753 | 0.1184 | 0.04245 | 105 | 6.83E-04 |
| rs1293752 | 0.1315 | 0.04717 | 105 | 3.11E-04 |
| rs1293751 | 0.1315 | 0.04717 | 105 | 3.11E-04 |
| rs2525848 | 0.1315 | 0.04717 | 105 | 3.11E-04 |
| rs1296061 | 0.1315 | 0.04717 | 105 | 3.11E-04 |
| rs2072137 | 0.4147 | 0.6038 | 61 | 4.23E-08 |
| rs1293749 | 0.134 | 0.05189 | 105 | 5.01E-04 |
| rs2003480 | 0.4367 | 0.283 | 96 | 8.95E-06 |
| rs1293748 | 0.134 | 0.05189 | 105 | 5.01E-04 |
| rs2240185 | 0.4834 | 0.6698 | 54 | 9.43E-08 |
| rs929291 | 0.1497 | 0.07547 | 104 | 2.70E-03 |
| rs1293747 | 0.2986 | 0.2217 | 99 | 1.58E-02 |
| rs1293746 | 0.285 | 0.2075 | 101 | 1.38E-02 |
| rs16942430 | 0.05797 | 0.01887 | 106 | 1.55E-02 |
| rs1293745 | 0.2858 | 0.2028 | 101 | 8.37E-03 |
| rs1293744 | 0.2858 | 0.2028 | 101 | 8.37E-03 |
| rs1293743 | 0.1613 | 0.06604 | 105 | 1.87E-04 |
| rs15895 | 0.1624 | 0.06604 | 105 | 1.63E-04 |
| rs574059077 | 0 | 0.00472 | 106 | 7.05E-08 |
| rs13311 | 0.4721 | 0.684 | 52 | 1.26E-09 |
| rs1058480 | 0.1626 | 0.06604 | 105 | 1.60E-04 |
| rs3815178 | 0.1884 | 0.1226 | 104 | 1.56E-02 |
| rs1859331 | 0.2756 | 0.1462 | 103 | 3.12E-05 |

| | | | | |
|---|---|---|---|---|
| **rs1859330** | 0.2762 | 0.1462 | 103 | 2.88E-05 |
| rs1859329 | 0.1884 | 0.1226 | 104 | 1.56E-02 |
| rs7299132 | 0.1884 | 0.1226 | 104 | 1.56E-02 |
| rs6489879 | 0.1884 | 0.1226 | 104 | 1.56E-02 |
| rs4238033 | 0.1884 | 0.1226 | 104 | 1.56E-02 |
| rs4767041 | 0.1954 | 0.1321 | 104 | 2.18E-02 |
| **rs10850102** | 0.1177 | 0.2642 | 98 | 1.65E-10 |
| rs7955267 | 0.1954 | 0.1321 | 104 | 2.18E-02 |
| rs7311182 | 0.188 | 0.1226 | 104 | 1.61E-02 |
| rs73433165 | 0.000162 | 0.004717 | 106 | 2.33E-04 |
| rs10735079 | 0.195 | 0.1274 | 104 | 1.41E-02 |
| rs6489880 | 0.1884 | 0.1226 | 104 | 1.56E-02 |
| rs7980275 | 0.1949 | 0.1321 | 104 | 2.27E-02 |
| rs7977345 | 0.1954 | 0.1321 | 104 | 2.18E-02 |
| rs6489881 | 0.1954 | 0.1321 | 104 | 2.18E-02 |
| rs6489882 | 0.1884 | 0.1226 | 104 | 1.56E-02 |
| rs7131998 | 0.1853 | 0.1274 | 104 | 3.21E-02 |
| rs2269899 | 0.1924 | 0.1368 | 104 | 4.25E-02 |
| rs11066456 | 0.1452 | 0.2075 | 93 | 1.17E-02 |
| rs12427406 | 0.126 | 0.1887 | 95 | 7.27E-03 |
| rs10850104 | 0.3358 | 0.4292 | 83 | 4.72E-03 |
| rs11066457 | 0.3688 | 0.4528 | 79 | 1.28E-02 |
| rs34647135 | 0.3358 | 0.4292 | 83 | 4.72E-03 |
| rs7965570 | 0.1332 | 0.2123 | 94 | 9.47E-04 |
| rs1974518 | 0.4003 | 0.4764 | 77 | 2.63E-02 |
| rs71465868 | 0.0328 | 0 | 106 | 7.36E-03 |
| rs12824584 | 0.00065 | 0.004717 | 106 | 3.76E-02 |
| rs146042277 | 0.000487 | 0.004717 | 106 | 1.56E-02 |
| rs10850105 | 0.1475 | 0.08962 | 104 | 1.89E-02 |
| rs7310667 | 0.1665 | 0.1132 | 104 | 3.99E-02 |
| rs4767042 | 0.4003 | 0.4764 | 77 | 2.63E-02 |
| rs78069989 | 0.00341 | 0.01887 | 106 | 4.01E-04 |
| rs78220999 | 0.1468 | 0.08962 | 104 | 2.00E-02 |
| rs4435062 | 0.3709 | 0.4481 | 82 | 2.23E-02 |
| rs150390228 | 0.1468 | 0.08962 | 104 | 2.00E-02 |
| rs10744789 | 0.1468 | 0.08962 | 104 | 2.00E-02 |
| rs140303054 | 0.000487 | 0.004717 | 106 | 1.56E-02 |
| rs57771566 | 0.002923 | 0.01415 | 106 | 5.05E-03 |
| rs138971703 | 0.002436 | 0.01415 | 106 | 1.58E-03 |
| rs4238034 | 0.147 | 0.08962 | 104 | 1.98E-02 |
| rs4766677 | 0.4021 | 0.4764 | 77 | 3.01E-02 |
| rs4766678 | 0.4029 | 0.4858 | 77 | 1.56E-02 |
| rs58603713 | 0.002923 | 0.01415 | 106 | 5.05E-03 |
| rs2158393 | 0.3707 | 0.4481 | 82 | 2.20E-02 |
| rs2072136 | 0.3704 | 0.4481 | 82 | 2.14E-02 |
| rs2072135 | 0.1466 | 0.2028 | 95 | 2.37E-02 |
| rs60439830 | 0.00341 | 0.01887 | 106 | 4.01E-04 |

| | | | | |
|---|---|---|---|---|
| rs55688670 | 0.03556 | 0.07075 | 103 | 7.40E-03 |
| rs45607836 | 0.03556 | 0.07075 | 103 | 7.40E-03 |
| rs4767044 | 0.1174 | 0.06132 | 104 | 1.20E-02 |
| rs2240189 | 0.1565 | 0.2123 | 96 | 2.89E-02 |
| rs1557866 | 0.1172 | 0.06132 | 104 | 1.22E-02 |
| rs45489899 | 0.00341 | 0.01887 | 106 | 4.01E-04 |
| rs3937434 | 0.1174 | 0.06132 | 104 | 1.20E-02 |
| rs2016831 | 0.1174 | 0.06132 | 104 | 1.20E-02 |
| rs11837165 | 0.000325 | 0.004717 | 106 | 3.75E-03 |
| rs757405 | 0.1179 | 0.0566 | 104 | 6.11E-03 |
| rs73422036 | 0.003248 | 0.01887 | 106 | 2.62E-04 |
| rs45620632 | 0.001624 | 0.0283 | 106 | 2.35E-14 |
| rs11837367 | 0.003248 | 0.01887 | 106 | 2.62E-04 |
| rs2010604 | 0.1387 | 0.08491 | 104 | 2.51E-02 |
| rs739903 | 0.1772 | 0.2358 | 95 | 2.85E-02 |
| rs2072133 | 0.195 | 0.2594 | 93 | 2.05E-02 |
| rs4767045 | 0.1181 | 0.05189 | 104 | 3.08E-03 |
| rs45583340 | 0.000162 | 0.004717 | 106 | 2.33E-04 |
| rs75755877 | 0.002923 | 0.01415 | 106 | 5.05E-03 |
| rs10744791 | 0.1181 | 0.05189 | 104 | 3.08E-03 |
| rs73422042 | 0.002923 | 0.01415 | 106 | 5.05E-03 |
| rs7023 | 0.1772 | 0.2358 | 95 | 2.85E-02 |
| rs2251109 | 0.2369 | 0.3538 | 84 | 9.15E-05 |
| rs116082988 | 0 | 0.00472 | 106 | 7.05E-08 |
| rs17078308 | 0.004385 | 0.01887 | 106 | 2.89E-03 |
| rs144151884 | 0 | 0.00472 | 106 | 7.05E-08 |
| rs116590098 | 0 | 0.00472 | 106 | 7.05E-08 |
| rs2531750 | 0.2363 | 0.1509 | 104 | 3.88E-03 |
| rs7621856 | 0.4756 | 0.3821 | 87 | 7.30E-03 |
| rs2286489 | 0.448 | 0.6085 | 60 | 3.95E-06 |
| rs113497906 | 0.000162 | 0.004717 | 106 | 2.33E-04 |
| rs6770261 | 0.2834 | 0.3585 | 85 | 1.73E-02 |
| rs7634267 | 0.02079 | 0.08019 | 104 | 1.19E-08 |
| rs9857669 | 0.1639 | 0.2264 | 92 | 1.61E-02 |
| rs7641997 | 0.1655 | 0.1132 | 102 | 4.32E-02 |
| rs575208313 | 0.2441 | 0.316 | 89 | 1.68E-02 |
| rs143341618 | 0.000162 | 0.004717 | 106 | 2.33E-04 |
| rs567829326 | 0 | 0.00472 | 106 | 7.05E-08 |
| rs62242259 | 0.1807 | 0.09434 | 102 | 1.22E-03 |
| rs9867918 | 0.1622 | 0.217 | 93 | 3.43E-02 |
| rs186253736 | 0.000162 | 0.004717 | 106 | 2.33E-04 |
| rs74850924 | 0.001299 | 0.01415 | 106 | 9.37E-06 |
| rs369845989 | 0.00065 | 0.004717 | 106 | 3.76E-02 |
| rs4327428 | 0.1468 | 0.2358 | 90 | 3.54E-04 |
| **rs2108917** | 0.2746 | 0.5142 | 74 | 2.61E-14 |
| rs720626 | 0.01932 | 0.04717 | 104 | 4.66E-03 |

| | | | | |
|---|---|---|---|---|
| rs6768156 | 0.2813 | 0.5047 | 76 | 1.67E-12 |
| rs189427751 | 0.000162 | 0.004717 | 106 | 2.33E-04 |
| rs758388 | 0.01965 | 0.0566 | 103 | 2.16E-04 |
| rs2077017 | 0.01965 | 0.06132 | 103 | 3.22E-05 |
| rs9818982 | 0.1504 | 0.2075 | 93 | 2.27E-02 |
| rs17279437 | 0.05196 | 0.1226 | 95 | 8.03E-06 |
| rs1468541 | 0.02371 | 0.09906 | 102 | 1.47E-11 |
| rs565220072 | 0.000162 | 0.004717 | 106 | 2.33E-04 |
| rs13314717 | 0.003735 | 0.0283 | 106 | 1.75E-07 |
| rs17078335 | 0.000812 | 0.009434 | 106 | 1.95E-04 |
| rs57133084 | 0.004547 | 0.01415 | 106 | 4.82E-02 |
| **rs2742396** | 0.4449 | 0.2123 | 97 | 1.88E-11 |
| **rs2531748** | 0.4591 | 0.2264 | 97 | 2.18E-11 |
| rs6771661 | 0.001462 | 0.009434 | 106 | 5.98E-03 |
| rs59375543 | 0.000487 | 0.004717 | 106 | 1.56E-02 |
| rs2252547 | 0.3509 | 0.5283 | 74 | 1.16E-07 |
| rs543762608 | 0.00065 | 0.004717 | 106 | 3.76E-02 |
| rs12493913 | 0.3553 | 0.25 | 98 | 1.59E-03 |
| rs576940167 | 0.00065 | 0.004717 | 106 | 3.76E-02 |
| rs17078339 | 0.3115 | 0.1981 | 101 | 4.38E-04 |
| rs34987516 | 0.3284 | 0.2075 | 101 | 2.20E-04 |
| rs142086756 | 0.000325 | 0.004717 | 106 | 3.75E-03 |
| rs184263104 | 0.000325 | 0.004717 | 106 | 3.75E-03 |
| rs2531747 | 0.1843 | 0.3349 | 92 | 3.74E-08 |
| rs9848415 | 0.302 | 0.1887 | 102 | 3.89E-04 |
| rs57126329 | 0.2267 | 0.09906 | 105 | 1.12E-05 |
| rs2159272 | 0.2808 | 0.467 | 78 | 3.79E-09 |
| rs7644870 | 0.3404 | 0.2075 | 101 | 5.68E-05 |
| rs1860263 | 0.3454 | 0.2123 | 101 | 5.80E-05 |
| rs545193808 | 0 | 0.00472 | 106 | 7.05E-08 |
| rs13064991 | 0.2272 | 0.1415 | 101 | 3.28E-03 |
| rs28437706 | 0.06382 | 0.1038 | 101 | 2.05E-02 |
| rs9852457 | 0.06382 | 0.1038 | 101 | 2.05E-02 |
| rs182605899 | 0.000325 | 0.004717 | 106 | 3.75E-03 |
| rs59776512 | 0.06285 | 0.1179 | 100 | 1.35E-03 |
| rs73062389 | 0.004385 | 0.01415 | 106 | 4.11E-02 |
| rs7615978 | 0.06479 | 0.1179 | 100 | 2.28E-03 |
| rs543563855 | 0 | 0.00472 | 106 | 7.05E-08 |
| rs7618553 | 0.06479 | 0.1038 | 101 | 2.47E-02 |
| rs147310206 | 0.000325 | 0.004717 | 106 | 3.75E-03 |
| rs188376831 | 0 | 0.00472 | 106 | 7.05E-08 |
| rs2271616 | 0.01153 | 0.0283 | 106 | 2.80E-02 |
| rs11085726 | 0.000325 | 0.004717 | 106 | 3.75E-03 |
| rs2304256 | 0.2061 | 0.316 | 85 | 1.10E-04 |
| rs12720270 | 0.1916 | 0.283 | 89 | 9.51E-04 |
| rs34725611 | 0.2061 | 0.3019 | 87 | 7.46E-04 |

| | | | | |
|---|---|---|---|---|
| rs569826524 | 0.4432 | 0.6038 | 59 | 3.78E-06 |
| rs12610298 | 0.1892 | 0.25 | 93 | 2.68E-02 |
| rs62130729 | 0.189 | 0.2594 | 91 | 1.04E-02 |
| rs280499 | 0.07584 | 0.1179 | 99 | 2.40E-02 |
| rs280500 | 0.03832 | 0.1132 | 99 | 6.03E-08 |
| rs12720218 | 0.02793 | 0.08962 | 99 | 2.24E-07 |
| rs280501 | 0.04206 | 0.1132 | 99 | 7.68E-07 |
| rs71327006 | 0.04726 | 0.009434 | 106 | 9.75E-03 |
| rs547178387 | 0.000487 | 0.004717 | 106 | 1.56E-02 |
| rs559851604 | 0.000487 | 0.004717 | 106 | 1.56E-02 |
| rs71327007 | 0.04726 | 0.009434 | 106 | 9.75E-03 |
